# Ownership and Quality of Primary Care: A Comparative Analysis of Public and Faith-Based Providers in Malawi

**DOI:** 10.1101/2025.11.18.25340538

**Authors:** Wiktoria Tafesse, Precious Chitsulo, Bingling She, Joseph H. Collins, Mariana Suarez, Dominic Nkhoma, Luigi Siciliani, Martin Chalkley, Sakshi Mohan, Watipaso Mulwafu, Emmanuel Mnjowe, Timothy B. Hallett, Joseph Mfutso-Bengo, Timothy Colbourn

**Affiliations:** Centre for Health Economics, University of York, York, England, United Kingdom; Health Economics and Policy Unit, Kamuzu University of Health Sciences, Lilongwe, Malawi; Department of Infectious Disease Epidemiology, School of Public Health, Imperial College London, London, UK; Department of Economics and Related Studies, University of York, York, England, United Kingdom; University College London Global Business School for Health London, London, England, United King-dom; Institute for Global Health, University College London, London, England, United Kingdom

**Author notes:** Corresponding author: Wiktoria Tafesse.

## Abstract

Non-profit, faith-based providers (FBPs) play a major role in primary care delivery across Sub-Saharan Africa and are increasingly integrated into national health systems, yet evidence on how the quality of faith-based primary care compares to that of public providers remains limited. Using healthcare worker observations, patient exit interviews, and patient follow-up data from Malawi, we compare consultation duration, clinical content, and patient-reported outcomes across faith-based and government facilities. Ordinary least squares estimates controlling for facility-, provider-, and shift-level characteristics, as well as patient health conditions, indicate that FBPs conduct significantly longer consultations, by around 1.9 minutes (75% of the sample mean), and perform a greater number of key clinical processes, including physical examinations and diagnostic tests. Patient-reported data corroborate these findings. Patients attending FBPs report receiving more tests and examinations and, at follow-up, are more likely to state that their treatment is working. These findings provide new evidence that publicly supported FBPs deliver higher-quality primary care on both process measures and patient-reported outcomes, underscoring the importance of accounting for provider ownership when examining variation in access to quality primary care in Sub-Saharan Africa.

## 1 Introduction

Access to high quality primary healthcare remains a persistent challenge across low- and middle-income countries (LMICs) (Das and Hammer, 2014; Kruk et al., 2018). Beyond the scarcity of healthcare resources, a growing body of evidence documents notably short consultation times, often well below the WHO ten minute recommendation, and inadequate quality of care during patient interactions, both of which have adverse consequences for health outcomes (Das et al., 2008; Leonard and Masatu, 2010; Das et al., 2012; Das and Hammer, 2014; Das et al., 2016; Irving et al., 2017; Rowe et al., 2018; Okeke, 2021; Kovacs and Lagarde, 2022; King et al., 2023; Banerjee et al., 2023; Sheffel et al., 2024; Daniels et al., 2025). Healthcare markets in the Global South are further characterised by a substantial presence of non-governmental providers and a sparse distribution of facilities, limiting provider choice, particularly in rural areas (Grépin, 2016; Sriram et al., 2024). While economic theory suggests several mechanisms that may generate variation in healthcare performance by facility ownership (Sloan, 2000), robust empirical evidence remains limited across LMICs (Basu et al., 2012; Herrera et al., 2014; Coarasa et al., 2017).

Non-profit faith-based providers (FBPs) constitute the largest non-governmental providers in Sub-Saharan Africa (SSA), where they deliver between 30% and 70% of healthcare services (Olivier et al., 2015) Due to fiscal constraints that limit public sector expansion, FBPs are often integrated into national health systems through contracting arrangements and salary support (Watson et al., 2016; Duff and Buckingham, 2015; Whyle and Olivier, 2017; Boulenger et al., 2014; Olivier et al., 2015; Manthalu et al., 2016; Tafesse et al., 2019). Yet, it remains unclear whether FBPs should be regarded as complementary to public healthcare provision. Using data from direct healthcare worker observations and patient exit interviews this study provides new evidence on differences in the quality of primary care across faith-based and public primary care facilities in Malawi. We analyse variation in outpatient consultation duration and the content of consultations, which are frequently used intermediary measures of quality across (see Das et al. (2008); Das and Hammer (2014); Das et al. (2016); Irving et al. (2017); Okeke (2021)). We further examine differences in patient reported outcomes at follow-up, comparing patients who visited differently managed facilities.

OLS regressions controlling for facility-, healthcare worker-, shift-level covariates as well as the patients’ health condition and the order of being seen by the provider, establish that healthcare workers in faith-based facilities spend on average approximately 1.9 more minutes per consultation, corresponding to 75% of the sample consultation duration average, and are more likely to have performed a physical examination, diagnostics and monitoring activities and prescribed medication, compared to their government counterparts. Our results are robust to controlling for patient case-mix, patient order and daily caseload. Our data collection design mitigates a potential Hawthorne effect where healthcare observations may induce better healthcare worker performance. Observations began unannounced and continued over several shifts, reducing anticipation and allowing any Hawthorne effect to dissipate, as prior evidence indicates it typically disappears after as few as ten consultations (Leonard and Masatu, 2006). Moreover, we show that differential Hawthorne effects by management are not driving our findings.

The findings on the contents of the consultations using direct healthcare worker observations are largely confirmed from analysing patient self-reported data which also allows us to account for variation in patient characteristics, including self-reported severity of illness. Patients attending outpatient consultations at faith-based primary health facilities are more likely to report having had a physical examination, receiving at least one test and are more likely to report having been diagnosed with a new condition. At follow-up, patients who visited FBPs were more likely to report that the treatment which they received is working, relative to patients visiting government facilities. As such, this study presents comprehensive support in favour of FBPs delivering higher quality of care measured by intermediate process measures as well as patient reported outcomes.

Our study contributes to the limited evidence on differences in the quality of primary care and health outcomes when comparing public providers with the most frequently publicly supported non-governmental healthcare networks in SSA, faith-based organisations. Although existing literature documents differences in quality of care between private-for-profit and public providers, private-for-profit and non-profit non-governmental providers, public and non-public facilities, or across decentralised organisational arrangements (Leonard et al., 2007; Leonard and Masatu, 2007, 2010; Das et al., 2016; Daniels et al., 2022; King et al., 2023; Su et al., 2021), far less is known about FBPs specifically. FBPs are perceived to deliver higher-quality care and to generate greater patient satisfaction than public and other private providers in Africa, a perception often attributed to their stronger intrinsic and pro-poor motivation (Adongo et al., 2021; Bawuah et al., 2024; Olivier et al., 2015; Reinikka and Svensson, 2010; Bjorvatn and Svensson, 2016; Collins et al., 2025). However, patient satisfaction does not necessarily reflect true quality of care, as it is subject to social desirability bias.

Our results are consistent with Leonard and Masatu (2007) who show that healthcare workers at private and church-based health facilities had higher practice quality scores than those in public facilities. Moreover, we show that systematic differences in consultation quality by facility ownership are mirrored in patient-reported outcomes at follow-up. Our paper also adds to the broader understanding of primary health care provision in LMICs. Our study reveals a higher daily caseload per primary healthcare worker than estimates derived from retrospectively dividing reported monthly caseloads per facility by the number of staff (Sheffel et al., 2024; Kovacs and Lagarde, 2022; Daniels et al., 2025). We show that the average consultation time of 2.5 minutes per patient in Malawi is shorter than the frequently reported 4–9 minutes observed in other LMICs (Das et al., 2008, 2012; Okeke, 2021; Kovacs and Lagarde, 2022). Furthermore, we observe that a higher caseload is negatively associated with consultation time in contrast with existing evidence for SSA (Mæstad et al., 2010; Kovacs and Lagarde, 2022).

## 2 The Malawian Healthcare System

Malawi’s health system is organised in a three-tier structure of primary, secondary, and tertiary healthcare provision under the Government of Malawi Ministry of Health (Malawi Ministry of Health, 2023a). More than half of all health facilities in the country are publicly owned, while approximately 15 percent are non-profit FBPs managed under the umbrella organisation - the Christian Health Association of Malawi (CHAM). Although a substantial share of facilities are operated by private-for-profit entities, NGOs, and other non-profit organisations, the vast majority of these are clinics that deliver lower-level primary care. CHAM manages 197 health facilities and 11 medical training colleges across the country (Malawi Ministry of Health, 2023b; Christian Health Association of Malawi, 2025). Its facilities are present in all districts except one and are predominantly located in rural areas, serving an estimated 37 percent of the Malawian population and delivering up to 75 percent of health services in rural and hard-to-reach areas (Christian Health Association of Malawi, 2025).

Public facilities provide care free of charge at the point of use, whereas CHAM facilities levy user fees for services not covered under Service Level Agreements, which typically include maternal, newborn, and child health services (Malawi Ministry of Health, 2023a). Under these agreements, FBPs deliver agreed upon services at no cost to users, with the government reimbursing CHAM for the associated costs. Moreover, the Government of Malawi pays salaries for CHAM staff, and CHAM facilities are fully integrated into district health planning and referral systems.

Our study focuses on primary care provision and we restrict our analytical sample to health centres and community hospitals which are primary-level facilities found for both ownership types in our data. In Malawi, health centres are primary-level facilities providing outpatient and maternity services, intended to serve a catchment population of approximately 10,000 people. Community hospitals, often found in rural areas, with greater capacity than health centres, and provide outpatient and inpatient services (Ministry of Health, Malawi, 2017).

## 3 Data and Descriptive Statistics

### 3.1 Data Collection

We use cross-sectional data collected with the aim to characterise the Malawian health-care system as part of the Thanzi La Mawa (TLM) project.^1^ The data collection included all of the country’s five government owned Central Hospitals (national referral hospitals), and 24 facilities stratified by ownership (government/CHAM), level of care (primary/secondary), urban/rural location, and high/low catchment population. Three enumerators visited each facility and conducted a health facility audit, direct healthcare worker observations through a Time and Motion Study (TMS) and patient exit interviews between January and May 2024. Patients were also contacted by phone for a follow-up interview.

#### 3.1.1 Healthcare Worker Observations

Enumerators with a clinical background^2^ observed healthcare workers using a TMS methodology used to quantify how healthcare providers allocate their time across different tasks Kalne and Mehendale (2022). Healthcare workers were selected using a random sample based on the facility duty roster, aiming to obtain a representative mix of healthcare worker cadres. Smaller facilities were over-sampled to ensure that at least one clinician was observed when available. The data collection was designed to minimise potential bias in healthcare worker attendance and performance by not announcing the start of the healthcare worker observations and following the same healthcare worker during consecutive shifts (aiming for at least three day shifts or one night shift), which is likely to reduce anticipation and Hawthorne bias given evidence that it diminishes over time (Leonard and Masatu, 2006).

Each task performed by the healthcare worker was recorded with a start- and end-time to the nearest minute. Each activity, including patient-facing and non-patient-facing activities such as breaks, administrative tasks, attending meetings, and inactivity was recorded using 167 activity categories.^3^ Each new patient seen by the healthcare worker per shift was recorded and given a unique ID number. The health condition of the attending outpatients was recorded using a list of 21 health conditions. At the end of the first shift, a brief interview with the healthcare worker was conducted to gather information on their profession, gender, education, and years of experience in their current profession.

#### 3.1.2 Patient Interviews

Adults exiting a facility were randomly selected for a patient-exit interview out of earshot of facility staff. The number of patients selected was based on the estimated daily patient load, as determined from facility registers or waiting room attendance. Respondents were asked about their demographic and socio-economic characteristics, whether they sought care for themselves or accompanied someone, health condition, illness severity and experiences during the visit. Respondents were also asked for their phone numbers and were contacted for a follow-up phone interview after two weeks. The follow-up interview gathered information on patient outcomes after their visit, including whether the patient’s illness had resolved and whether the treatment administered was effective.

### 3.2 Descriptive Statistics

#### 3.2.1 Healthcare Worker Observations

In line with the previous literature on consultation times (Irving et al., 2017), we focus on healthcare workers observed during day shifts across any outpatient department, encompassing general- adult- and children’s outpatient departments. Our analytical healthcare worker observation sample includes 43 healthcare workers (26 FBPs and 17 government employees) at 19 primary care facilities (11 managed by CHAM and 8 publicly managed). In total, our sample consists of 83 observation days (see Table A1 in the Appendix for shift-level descriptive statistics). We note a large difference in daily caseloads by staff and ownership. On average, healthcare workers at faith-based facilities attended to 27.26 outpatients per day, whereas government employees saw an average of 80.70 patients per day. This systematic difference between non-governmental and governmental facilities has been reported previously using multi-country data for Sub-Saharan Africa, Sheffel et al. (2024) and Mæstad et al. (2010) for Tanzania. On the other hand, our data implies higher average daily patient volumes per healthcare worker than previously reported in similar settings across (Kovacs and Lagarde, 2022; Sheffel et al., 2024; Daniels et al., 2025)

We calculate consultation duration in minutes using the difference between the start of the first patient-facing activity and the end of the last activity for each patient encounter.^4^ For each patient interaction, we map the activities performed by the healthcare worker to key processes of a primary healthcare consultation broadly following Byrne and Long (1976); Okeke (2021). We generate binary variables denoting whether the healthcare worker conducted any: physical examination, diagnostic/monitoring activity, or prescribed or dispensed any medication. Physical examination captures all direct clinical assessments conducted by the health worker, including measurement of vital signs and other hands-on examinations such as skin or eye assessment. Diagnostic and monitoring activities include all laboratory and imaging investigations, e.g. blood tests, urine tests, pathology investigations, imaging procedures, and monitoring tasks such as sample preparation or running tests. Prescribing and dispensing drugs comprises activities related to the provision or administration of medications, including prescribing, checking and dispensing prescriptions, preparing drugs, and administering oral, inhaled, intravenous, intramuscular, suppository, eye, or nasogastric drugs and fluids.

Table 1 displays the descriptive statistics for the analytical sample of observed outpatient consultations. 34 percent of consultations took place in religiously managed facilities, with the remainder in publicly managed facilities. The average consultation length is 2.52 minutes, which is shorter than what has been documented in other LMIC settings. For comparison, previous studies report mean consultation durations of 3.8 minutes in Delhi (Das and Sohnesen, 2007), 12.6 minutes across primary rural facilities in Senegal (Kovacs and Lagarde, 2022), and approximately 9 minutes in public-sector outpatient consultations in Nigeria (Okeke, 2021). Physical examinations were conducted in 11 percent of visits and diagnostic or monitoring activities in 4 percent. Healthcare workers dispensed or prescribed medication during 36 percent of consultations which is substantially lower than the 90 percent prescription rate reported in Senegal by Kovacs and Lagarde (2022). These figures suggest that a large share of patient encounters involve limited clinical assessment, consistent with earlier findings on provider effort in primary care settings Das et al. (2008).

**Table 1:** Descriptive statistics: Consultation observations.

|  | Mean | SD |
| --- | --- | --- |
| FBP consultation | 0.34 | 0.47 |
| Consultation time (minutes) | 2.52 | 4.36 |
| Consultation content: Physical examination | 0.11 | 0.31 |
| Consultation content: Diagnostics/Monitoring | 0.04 | 0.19 |
| Consultation content: Drug dispensing/prescribing | 0.36 | 0.48 |
| Day of healthcare worker observation | 1.92 | 0.82 |
| Healthcare worker observed during whole shift | 0.92 | 0.28 |
| Normal working day | 0.43 | 0.50 |
| Survey month: January | 0.09 | 0.29 |
| Survey month: February | 0.20 | 0.40 |
| Survey month: March | 0.24 | 0.43 |
| Survey month: April | 0.46 | 0.50 |
| Facility location: Rural | 0.55 | 0.50 |
| Facility type: Community Hospital | 0.68 | 0.46 |
| Facility type: Health Centre | 0.32 | 0.46 |
| Facility catchment population (1000) | 39.76 | 28.24 |
| Male healthcare worker | 0.61 | 0.49 |
| Healthcare worker professional experience (years) | 6.80 | 5.43 |
| Healthcare worker profession: Clinical Officer/Technician | 0.43 | 0.50 |
| Healthcare worker profession: Medical Assistant | 0.50 | 0.50 |
| Healthcare worker profession: Nurse Officer/Midwife Technician | 0.07 | 0.26 |
| Patient order | 46.18 | 43.01 |
| Health condition: Malaria | 0.42 | 0.49 |
| Health condition: ALRI | 0.18 | 0.39 |
| Health condition: ANC | 0.01 | 0.12 |
| Health condition: Diarrhoea | 0.04 | 0.20 |
| Health condition: Maternal/Newborn healthcare | 0.01 | 0.12 |
| Health condition: NCDs (incl. Mental Health & Other NCD) | 0.10 | 0.30 |
| Health condition: Other | 0.23 | 0.42 |
| Observations | 3905 |  |

*Notes:* This table reports descriptive statistics for observed healthcare-worker consultations conducted in outpatient departments of community hospitals and health centres included in the Thanzi La Mawa (TLM) project Time and Motion Study (TMS). Mean/proportion and standard deviation (SD) values for consultations in the analytical sample are presented. FBP is a binary indicator for whether the consultation was observed at a faith-based (CHAM) facility rather than a government facility. ALRI = acute lower respiratory infection; ANC = antenatal care; NCDs = non-communicable diseases. Patient order refers to the order of the consultation within the observed shift.

Around half of all consultations occurred in rural areas. Around two thirds of patient interactions were observed in community hospitals, with the rest in smaller health centres. In terms of provider characteristics, 43 percent of consultations were conducted by clinical officers or technicians, 50 percent by medical assistants, and 7 percent by nurse officers or midwife technicians. This distribution reflects the staffing composition of Malawi’s primary healthcare system, where most outpatient services are delivered by mid-level cadres rather than physicians.

Enumerators typically observed healthcare workers for nearly two full working days, and 92 percent of these observations covered the provider’s entire shift. Given these observation patterns, Hawthorne effects are unlikely to pose a significant threat to our analysis, as providers typically revert to routine performance after the first 10–15 consultations (Leonard and Masatu, 2006). 43 percent of observations occurred on normal working days, i.e., not during weekends or holidays. The case-mix of consultations is dominated by malaria (42%) potentially due to data collection during the wet season in Malawi. Acute lower respiratory infections (ALRI) including pneumonia, non-communicable diseases (NCDs) and diarrhoea comprise 18, 10 and 4 percent of all observed health conditions, respectively.

Summary statistics for observed consultations by facility management show that patient interactions with FBPs, compared to government providers, on average last twice as long (3.8 vs. 1.9 minutes) and are more likely to include physical examinations (21% vs. 5%) or diagnostic tests (7% vs. 2%) and dispensing or prescribing of drugs (50% vs. 29%), see Table A2 in the Appendix. A higher proportion of observed consultations are with clinical officers or nurse-midwives at CHAM facilities whereas medical assistants are more likely to be observed at government facilities. While the distribution of consultations with FBPs is similar across CHAM community hospitals and CHAM health centres, the analytical sample for government consultations comprises of a higher proportion of consultations at community hospitals compared to health centres. The average patient consultation order varies substantially across ownership, with mean values of 21.47 and 59.12 for faith-based and public consultations, respectively, reflecting the heterogeneity in daily caseload. Malaria constitutes nearly half of all observed health conditions among consultations at public facilities compared to 28 percent at FBPs. There is also a slightly higher proportion of patients presenting with ALRI at public facilities (20%) compared to at religiously managed facilities (16%). While the proportion of patients presenting with NCDs and diarrhoea are comparable across facility ownership, a larger share of patients seek maternal and newborn healthcare, including antenatal care (ANC) and other health conditions in faith-based facilities.

#### 3.2.2 Patient Reported Outcomes

For the analyses of the patient-level data, we restrict the analytical sample to individuals who sought care for themselves due to illness or accompanied someone who was unwell, at outpatient departments at community hospitals and health centres managed by CHAM and the government. Our empirical sample consists of 17 facilities in total of which 11 are managed by CHAM and 6 by the government. Table 2 presents the descriptive statistics for the analytical sample of clients exiting a primary care facility. More than two-thirds of all patients visited a FBP. Overall, a higher proportion of patients reported having received a physical examination (51%) compared with the findings from the direct healthcare worker observations. Although we do not have corresponding information on diagnostic or monitoring activities and drug prescription from the patient exit interview, 65 percent of patients reported receiving a test, 13 percent answered that they have been diagnosed with a new condition, and 90 percent reported having received some form of treatment or medication.

**Table 2:** Descriptive statistics: Patient exit interviews.

|  | Mean | SD |
| --- | --- | --- |
| Patient visited a FBP | 0.69 | 0.46 |
| Patient reported receiving: Physical examination | 0.51 | 0.50 |
| Patient reported receiving: Any test | 0.65 | 0.48 |
| Patient reported receiving: New diagnosis | 0.13 | 0.33 |
| Patient reported receiving: Any treatment/medication | 0.90 | 0.30 |
| Facility location: Rural | 0.50 | 0.50 |
| Facility type: Community hospital | 0.54 | 0.50 |
| Patient age: 0–5 | 0.36 | 0.48 |
| Patient age: 6–12 | 0.10 | 0.30 |
| Patient age: 13–17 | 0.07 | 0.25 |
| Patient age: 18–25 | 0.14 | 0.34 |
| Patient age: 26–49 | 0.24 | 0.43 |
| Patient age: 50+ | 0.09 | 0.29 |
| Male patient | 0.43 | 0.50 |
| Patient sought care for self | 0.50 | 0.50 |
| Household wealth quintile: Poorest | 0.31 | 0.46 |
| Household wealth quintile: Poor | 0.17 | 0.38 |
| Household wealth quintile: Middle | 0.18 | 0.38 |
| Household wealth quintile: Richer | 0.16 | 0.37 |
| Household wealth quintile: Richest | 0.17 | 0.37 |
| Health condition: Malaria | 0.27 | 0.44 |
| Health condition: ALRI | 0.18 | 0.38 |
| Health condition: Diarrhoea | 0.05 | 0.23 |
| Health condition: NCDs | 0.09 | 0.29 |
| Health condition: Other | 0.41 | 0.49 |
| Health status: Healthy | 0.04 | 0.20 |
| Health status: Mild symptoms | 0.52 | 0.50 |
| Health status: Moderate symptoms | 0.39 | 0.49 |
| Health status: Severe/life-threatening symptoms | 0.06 | 0.23 |
| Observations | 1118 |  |

*Notes:* This table reports descriptive statistics for adults sampled for a patient-exit interview as part of the Thanzi La Mawa (TLM) project data collection as they were leaving community hospitals and health centres and reported attending an outpatient department. Mean/proportion and standard deviation (SD) of patients in the analytical sample are reported. FBP is a binary variable indicating whether the patient attended a faith-based (CHAM) facility rather than a government facility. “Patient sought care for self” takes the value 1 if the respondent reported seeking care for themselves and 0 if they accompanied someone seeking care. Wealth quintiles are constructed using household asset ownership. Illness severity categories reflect patients’ self-reported symptoms at the time of exit. ALRI = acute lower respiratory infection; NCD = non-communicable disease.

The patient-level data provides further information on patient characteristics attending primary care outpatient consultations compared to data from the healthcare worker observations. Among all patients, 36 percent are children under five, and 24 percent are adults aged 26–49, followed by adults aged 18–25. There are more female (57%) than male (43%) patients. Half of respondents reported seeking care for themselves, as opposed to accompanying someone who was unwell. Almost one-third of all patients come from households in the lowest wealth quintile, while the remaining distribution across wealth quintiles is comparatively even. Self-reported conditions indicate that malaria is the most common health problem, followed by ALRI, NCDs, and diarrhoea.^5^ When asked about the severity of their symptoms upon exiting the facility, approximately half of all respondents reported having mild symptoms, 39 percent had moderate symptoms, and 6 percent had severe or life-threatening symptoms.

While there is a similar proportion of patients attending both facility ownerships who report receiving any treatment or medication, patients exiting CHAM primary care facilities report higher rates of physical examinations, diagnostic testing and receiving a new diagnosis, compared to those who attended consultations at public facilities (Appendix Table A3). A substantially higher proportion of CHAM patients attended community hospitals, whereas public-sector patients more commonly visited health centres. Children aged 0–5 constitute a larger share of FBP clients, while public facilities see relatively more adolescents and adults. Socio-economic differences are also pronounced. 40 percent of patients at public facilities belong to the poorest wealth quintile compared with 27 percent among CHAM clients. 22 percent of FBP patients are from the richest quintile versus only 5 percent in public facilities. Presenting health conditions are largely comparable across ownership types, though patients having attended a consultation at faith-based facilities report a slightly higher share of NCDs. Severity profiles are comparable as 45 percent of FBP patients and 43 percent of public-facility patients report moderate or severe/life-threatening symptoms.

Descriptive statistics for patients that were successfully contacted for a phone interview at follow-up are presented in Table 3. Compared to the full sample of interviewed outpatients exiting community hospitals and health centres, a slightly higher proportion of patients who attended FBPs were re-contacted at follow-up. The vast majority of all patients report that their sickness has been resolved (91%) and that the administered treatment is working (95%). Compared to the patients interviewed at baseline, a larger proportion of patients at follow-up come from non-poor household, potentially reflecting individuals who enumerators were able to contact by phone. Otherwise, the proportion of respondents who had moderate or severe symptoms upon exiting the facility, compared to mild or healthy symptoms, and the frequency of health conditions is comparable to the analytical patient sample upon facility exit. Summary statistics by facility management show that patients who visited CHAM facilities report slightly higher rates of sickness resolution (92% vs. 88%) and perceptions that treatment is working (96% vs. 93%) compared to patients attending public facilities (see Table A4 in the Appendix).

**Table 3:** Descriptive statistics: Patient follow-up interviews.

|  | Mean | SD |
| --- | --- | --- |
| Patient visited a FBP | 0.75 | 0.43 |
| Sickness resolved | 0.91 | 0.28 |
| Treatment is working | 0.95 | 0.21 |
| Follow-up: <2 weeks | 0.06 | 0.24 |
| Follow-up: 2–3 weeks | 0.56 | 0.50 |
| Follow-up: >3 weeks | 0.37 | 0.48 |
| Facility location: Rural | 0.51 | 0.50 |
| Facility type: Community hospital | 0.55 | 0.50 |
| Patient age: 0–5 | 0.37 | 0.48 |
| Patient age: 6–12 | 0.12 | 0.32 |
| Patient age: 13–17 | 0.05 | 0.22 |
| Patient age: 18–25 | 0.13 | 0.34 |
| Patient age: 26–49 | 0.25 | 0.44 |
| Patient age: 50+ | 0.08 | 0.27 |
| Male patient | 0.45 | 0.50 |
| Patient sought care for self | 0.47 | 0.50 |
| Household wealth quintile: Poorest | 0.17 | 0.37 |
| Household wealth quintile: Poor | 0.21 | 0.40 |
| Household wealth quintile: Middle | 0.20 | 0.40 |
| Household wealth quintile: Richer | 0.20 | 0.40 |
| Household wealth quintile: Richest | 0.23 | 0.42 |
| Health condition: Malaria | 0.26 | 0.44 |
| Health condition: ALRI | 0.19 | 0.40 |
| Health condition: Diarrhoea | 0.05 | 0.23 |
| Health condition: NCDs | 0.09 | 0.29 |
| Health condition: Other | 0.40 | 0.49 |
| Health status: Healthy | 0.02 | 0.14 |
| Health status: Mild symptoms | 0.52 | 0.50 |
| Health status: Moderate symptoms | 0.41 | 0.49 |
| Health status: Severe/life-threatening symptoms | 0.05 | 0.23 |
| Observations | 675 |  |

*Notes:* This table reports descriptive statistics for adults who were re-contacted by phone after two weeks or more after being included in the Thanzi La Mawa (TLM) patient-exit interviews as they were leaving community hospitals and health centres and reported attending an outpatient department. Mean/proportion and standard deviation (SD) of patients in the analytical sample are reported. All variables besides “Sickness resolved”, “Treatment is working” and Follow-up duration were collected during the patient exit interview. FBP is a binary variable indicating whether the patient attended a faith-based (CHAM) facility rather than a government facility. “Patient sought care for self” takes the value 1 if the respondent reported seeking care for themselves and 0 if they accompanied someone seeking care. Wealth quintiles are constructed using household asset ownership. Illness severity categories reflect patients’ self-reported symptoms at the time of exit. ALRI = acute lower respiratory infection; NCD = non-communicable disease.

## 4 Empirical Strategy

We estimate the following regression model using OLS as displayed in Equation 1.

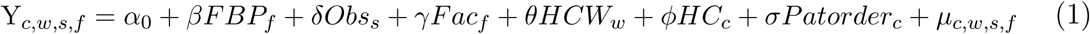

For analyses using data on direct healthcare observations, the outcome variable *Y* are either consultation duration or the likelihood that a key activity (physical examination, diagnostics/monitoring or drug dispensing/prescribing) is conducted during consultation *c*, for healthcare worker *w*, during shift *s*, in facility *f*. Our main explanatory variable of interest is *FBP_f_*, a binary indicator taking the value 1 if the healthcare worker is employed at a faith-based primary care facility managed by CHAM, and 0 if they work at a government-managed facility.

*Obs_s_* is a vector of covariates capturing variation in data collection. Specifically, we control for the day of observation for a given healthcare worker, whether the clinician was observed for the full duration of their shift, survey month, and whether the observation took place on a normal working day versus a weekend or public holiday.

We also include facility characteristics, denoted *Fac_f_*, which comprise the rural or urban location of the facility, whether the facility is a community hospital relative to a health centre, and the facility’s catchment population. *HCW_w_* denotes healthcare worker characteristics, including cadre (clinical officer/technician, medical assistant, or nurse officer/midwife technician), years of experience, and gender. Finally, we account for the patient’s health condition, *HC_c_*, as well as the patient’s order of being attended to during the shift, *Patorder_c_*, given that provider effort has been shown to reduce with increasing patient order (Das and Sohnesen, 2006). Robust standard errors are used throughout. As the number of facilities is small (n=19), we report wild bootstrapped standard errors as a robustness check.

We employ similar regression models when conducting separate analyses at the patient level using the facility-exit and follow-up data, respectively. We estimate a linear probability model for patient *p* reporting having received a physical examination, any test, being diagnosed with a new condition or having received any treatment or medication after their healthcare visit. In addition to the facility-level covariates included in Equation 1, we also control for the patient’s age, household wealth quintile, whether the individual sought care for themselves or someone they accompanied, and self-reported symptom severity.^6^ These covariates are also included when estimating the relationship between facility management and patient reports of their illness having resolved or that the administered treatment is working at follow-up, with additional adjustment for the time since the patient exit interview.

## 5 Results

### 5.1 Healthcare Worker Observations

Table 4 presents the regression results of Equation 1 on consultation time. Sets of control variables are added sequentially. On average, FBPs spend 2.4 more minutes per outpatient interaction compared to public providers after accounting for observation day, whether the clinician’s full shift was observed, month of observation and whether the observation took place during a normal working day (column 1). Including facility-level covariates increases the magnitude of the coefficient on *FBP* to 2.9. In column 3, we additionally control for healthcare worker profession, professional experience and gender, yielding a coefficient of 2.5. The coefficient decreases to 2.2 after controlling for clients’ observed health condition (column 4). Accounting for patient order further reduces the estimated difference, indicating that healthcare workers in faith-based facilities spend approximately 1.9 more minutes per consultation compared to those in government facilities (column 5). This estimate corresponds to approximately 75% of the sample mean.

**Table 4:** Faith-based facility ownership and consultation time.

|  | Dependent variable: Consultation time (minutes) |  |  |  |  |
| --- | --- | --- | --- | --- | --- |
|  | (1) | (2) | (3) | (4) | (5) |
| FBP | 2.364***<br>(0.174) | 2.869***<br>(0.171) | 2.520***<br>(0.363) | 2.189***<br>(0.357) | 1.907***<br>(0.353) |
| Observation day 2 | -0.231<br>(0.169) | -0.300*<br>(0.177) | -0.277<br>(0.177) | -0.258<br>(0.176) | -0.081<br>(0.179) |
| Observation day 3+ | -0.400**<br>(0.180) | -0.287<br>(0.174) | -0.212<br>(0.178) | -0.178<br>(0.174) | -0.186<br>(0.173) |
| Healthcare worker observed whole shift | 1.042***<br>(0.236) | 0.625***<br>(0.235) | 0.366<br>(0.249) | 0.340<br>(0.250) | 0.465*<br>(0.249) |
| Survey month: February | 1.962***<br>(0.290) | 2.474***<br>(0.290) | 2.490***<br>(0.474) | 2.289***<br>(0.473) | 1.937***<br>(0.470) |
| Survey month: March | 1.836***<br>(0.275) | 1.799***<br>(0.348) | 1.075<br>(0.759) | 1.098<br>(0.756) | 0.978<br>(0.751) |
| Survey month: April | 1.887***<br>(0.241) | 1.454***<br>(0.328) | 1.015<br>(0.800) | 0.740<br>(0.814) | 0.367<br>(0.811) |
| Normal working day | 0.064<br>(0.145) | 0.103<br>(0.146) | 0.107<br>(0.146) | 0.120<br>(0.143) | 0.019<br>(0.142) |
| Facility location: Rural |  | 0.348**<br>(0.164) | 0.608*<br>(0.317) | 0.630**<br>(0.316) | 0.379<br>(0.317) |
| Facility type: Community Hospital |  | 1.055***<br>(0.288) | 1.600***<br>(0.456) | 1.778***<br>(0.492) | 1.999***<br>(0.492) |
| Facility catchment population (1000s) |  | 0.030***<br>(0.004) | 0.032***<br>(0.005) | 0.028***<br>(0.005) | 0.027***<br>(0.005) |
| Male healthcare worker |  |  | -0.218<br>(0.321) | -0.443<br>(0.321) | -0.312<br>(0.320) |
| Medical Assistant |  |  | 0.079<br>(0.271) | -0.033<br>(0.272) | -0.000<br>(0.271) |
| Nurse Officer/Midwife Technician |  |  | 1.566**<br>(0.713) | 2.139***<br>(0.729) | 1.854**<br>(0.726) |
| Professional experience (years) |  |  | -0.037**<br>(0.018) | -0.012<br>(0.018) | 0.012<br>(0.018) |
| Health condition: ALRI |  |  |  | 0.555***<br>(0.147) | 0.520***<br>(0.146) |
| Health condition: ANC |  |  |  | -2.323***<br>(0.706) | -2.535***<br>(0.710) |
| Health condition: Diarrhoea |  |  |  | 1.095***<br>(0.401) | 1.042***<br>(0.397) |
| Health condition: Maternal/Newborn healthcare |  |  |  | 1.034<br>(0.836) | 0.945<br>(0.823) |
| Health condition: NCDs |  |  |  | 2.503***<br>(0.320) | 2.391***<br>(0.316) |
| Health condition: Other |  |  |  | 1.667***<br>(0.194) | 1.596***<br>(0.192) |
| Patient order |  |  |  |  | -0.012***<br>(0.001) |
| Constant | -0.802*<br>(0.412) | -2.631***<br>(0.442) | -2.282***<br>(0.781) | -2.761***<br>(0.765) | -2.116***<br>(0.754) |
| Observations | 3905 | 3905 | 3905 | 3905 | 3905 |
| R<sup>2</sup> | 0.061 | 0.091 | 0.095 | 0.137 | 0.147 |

*Notes:* This table presents the results from Equation 1, using data on consultations from healthcare-worker observations in outpatient departments in community hospitals and health centres participating in the Thanzi La Mawa (TLM) Time and Motion Study. The outcome variable is consultation time (minutes). FBP equals 1 if the observed consultation was conducted at a faith-based (CHAM) facility and 0 if conducted at a government facility. The omitted reference category for observation day is Observation Day 1, for facility type it is health centre, for healthcare-worker profession it is Clinical Officer/Technician; and for health condition it is malaria. ALRI = acute lower respiratory infection, ANC= antenatal care, NCD = non-communicable disease. Robust standard errors are reported in parentheses. \* $p < 0.10$ , \*\* $p < 0.05$ , \*\*\* $p < 0.01$ .

The results provide several additional insights. Consultations are significantly longer in community hospitals compared to health centres and in facilities with larger catchment populations. Compared with clinical cadres, nursing and midwifery staff spend on average almost two minutes longer per patient. While ANC consultations are shorter than malaria consultations, other health conditions (ALRI, Diarrhoea and NCDs) take longer time. Patient order is negatively associated with consultation duration, with each subsequent patient seen during the day spending approximately 0.012 fewer minutes with their provider. These findings are consistent with Das and Sohnesen (2006) who report that provider effort, including consultation time, declines over time.

Robustness checks, presented in Table A5 in the Appendix, confirm the consistency of these findings. Restricting the analysis to providers who were observed for a full day shift (columns 1–2) and excluding unusually long consultations of 30 minutes or more following Okeke (2021) (columns 3–4) yields similar results. We also estimate models restricted to the second or later consecutive observed shift for each healthcare worker, drawing on evidence from Leonard and Masatu (2010) that the initial Hawthorne effect is typically larger among public-sector compared to non-governmental providers (columns 5–6). Analysing data for observation day 2-5 and controlling for the full set of covariates, suggest that FBPs spend on average 2.3 more minutes per outpatient consultation, com-pared to pubic providers. Finally, our results remain robust when using a wild cluster bootstrap for cluster-robust standard errors at the facility level in the presence of a small number of facilities.

In addition to longer consultations, staff in faith-based facilities are consistently more likely to conduct a physical examination, perform diagnostic or monitoring activities, and prescribe or dispense medication (see Tables 5, 6, and 7, respectively). The coefficients on *FBP* for both physical examinations and diagnostic or monitoring activities are sizeable, particularly when compared with the corresponding sample means of consultations in which these processes occur. Overall, our results align with Okeke (2021), who find a positive association between consultation length, the likelihood of conducting a physical examination, and the use of diagnostic tests, and Das and Sohnesen (2007) who demonstrate that extending a longer consultation duration increases checklist completion rates.

**Table 5:** Faith-based facility ownership and likelihood of healthcare workers conducting a physical examination.

|  | Dependent variable: Physical examination |  |  |  |  |
| --- | --- | --- | --- | --- | --- |
|  | (1) | (2) | (3) | (4) | (5) |
| FBP | 0.165***<br>(0.014) | 0.181***<br>(0.013) | 0.183***<br>(0.024) | 0.145***<br>(0.024) | 0.132***<br>(0.024) |
| Observation day 2 | 0.028**<br>(0.012) | 0.002<br>(0.012) | -0.000<br>(0.012) | 0.008<br>(0.012) | 0.016<br>(0.012) |
| Observation day 3+ | -0.005<br>(0.011) | -0.021*<br>(0.011) | -0.020*<br>(0.011) | -0.014<br>(0.011) | -0.014<br>(0.011) |
| Healthcare worker observed during whole shift | 0.043**<br>(0.020) | 0.001<br>(0.021) | -0.017<br>(0.022) | -0.004<br>(0.022) | 0.002<br>(0.022) |
| Survey month: February | 0.040*<br>(0.024) | 0.059**<br>(0.024) | 0.020<br>(0.033) | 0.005<br>(0.033) | -0.011<br>(0.033) |
| Survey month: March | 0.070***<br>(0.024) | 0.093***<br>(0.029) | 0.017<br>(0.047) | 0.040<br>(0.047) | 0.035<br>(0.047) |
| Survey month: April | 0.007<br>(0.022) | -0.013<br>(0.028) | -0.093*<br>(0.049) | -0.067<br>(0.049) | -0.084*<br>(0.049) |
| Normal working day | 0.039***<br>(0.010) | 0.038***<br>(0.010) | 0.023**<br>(0.010) | 0.021**<br>(0.010) | 0.017*<br>(0.010) |
| Facility location: Rural |  | 0.095***<br>(0.011) | 0.063***<br>(0.020) | 0.043**<br>(0.020) | 0.032<br>(0.020) |
| Facility type: Community Hospital |  | 0.060***<br>(0.021) | 0.112***<br>(0.027) | 0.062**<br>(0.028) | 0.073**<br>(0.028) |
| Facility catchment population (1000) |  | 0.001***<br>(0.000) | 0.002***<br>(0.000) | 0.001***<br>(0.000) | 0.001***<br>(0.000) |
| Male healthcare worker |  |  | 0.009<br>(0.018) | 0.025<br>(0.018) | 0.031*<br>(0.018) |
| Medical Assistant |  |  | 0.017<br>(0.019) | 0.009<br>(0.018) | 0.011<br>(0.018) |
| Nurse Officer/Midwife Technician |  |  | 0.102***<br>(0.039) | 0.038<br>(0.039) | 0.025<br>(0.040) |
| Professional experience (years) |  |  | 0.005***<br>(0.001) | 0.004***<br>(0.001) | 0.005***<br>(0.001) |
| Health condition: ALRI |  |  |  | 0.063***<br>(0.012) | 0.062***<br>(0.012) |
| Health condition: ANC |  |  |  | 0.465***<br>(0.063) | 0.456***<br>(0.062) |
| Health condition: Diarrhoea |  |  |  | 0.069***<br>(0.023) | 0.066***<br>(0.023) |
| Health condition: Maternal/Newborn healthcare |  |  |  | 0.042<br>(0.052) | 0.038<br>(0.051) |
| Health condition: NCDs |  |  |  | 0.086***<br>(0.019) | 0.081***<br>(0.019) |
| Health condition: Other |  |  |  | 0.096***<br>(0.014) | 0.093***<br>(0.013) |
| Patient order |  |  |  |  | -0.001***<br>(0.000) |
| Constant | -0.044<br>(0.031) | -0.144***<br>(0.033) | -0.142**<br>(0.055) | -0.141**<br>(0.055) | -0.112**<br>(0.055) |
| Observations | 3905 | 3905 | 3905 | 3905 | 3905 |
| R<sup>2</sup> | 0.080 | 0.107 | 0.117 | 0.155 | 0.159 |

*Notes:* This table presents the results from Equation 1, using data on consultations from healthcare-worker observations in outpatient departments in community hospitals and health centres participating in the Thanzi La Mawa (TLM) Time and Motion Study. The outcome variable is the likelihood of the healthcare worker being observed to conduct any type of physical examination during a patient consultation. FBP equals 1 if the observed consultation was conducted at a faith-based (CHAM) facility and 0 if conducted at a government facility. The omitted reference category for observation day is Observation Day 1, for facility type it is health centre, for healthcare-worker profession it is Clinical Officer/Technician; and for health condition it is malaria. ALRI = acute lower respiratory infection, ANC= antenatal care, NCD = non-communicable disease. Robust standard errors are reported in parentheses. \* $p < 0.10$ , \*\* $p < 0.05$ , \*\*\* $p < 0.01$ .

**Table 6:** Faith-based facility ownership and likelihood of healthcare workers conducting any diagnostics or monitoring of patients.

|  | Dependent variable: Diagnostics/Monitoring |  |  |  |  |
| --- | --- | --- | --- | --- | --- |
|  | (1) | (2) | (3) | (4) | (5) |
| FBP | 0.045***<br>(0.008) | 0.065***<br>(0.007) | 0.067***<br>(0.015) | 0.076***<br>(0.015) | 0.073***<br>(0.015) |
| Observation day 2 | -0.022***<br>(0.007) | -0.022***<br>(0.008) | -0.022***<br>(0.008) | -0.021***<br>(0.008) | -0.019**<br>(0.008) |
| Observation day 3+ | -0.036***<br>(0.008) | -0.034***<br>(0.008) | -0.029***<br>(0.007) | -0.029***<br>(0.008) | -0.029***<br>(0.008) |
| Observed during whole shift | 0.044***<br>(0.008) | 0.041***<br>(0.009) | 0.020**<br>(0.009) | 0.019*<br>(0.010) | 0.020**<br>(0.010) |
| Survey month: February | -0.028<br>(0.018) | 0.013<br>(0.018) | 0.050**<br>(0.022) | 0.055**<br>(0.022) | 0.051**<br>(0.022) |
| Survey month: March | -0.012<br>(0.017) | 0.052**<br>(0.023) | 0.063*<br>(0.035) | 0.061*<br>(0.035) | 0.059*<br>(0.035) |
| Survey month: April | -0.041***<br>(0.015) | 0.032<br>(0.021) | 0.072*<br>(0.039) | 0.072*<br>(0.040) | 0.068*<br>(0.040) |
| Normal working day | -0.027***<br>(0.007) | -0.030***<br>(0.006) | -0.030***<br>(0.007) | -0.029***<br>(0.007) | -0.030***<br>(0.007) |
| Facility location: Rural |  | 0.007<br>(0.006) | 0.051***<br>(0.015) | 0.054***<br>(0.015) | 0.051***<br>(0.015) |
| Facility type: Community Hospital |  | -0.054***<br>(0.015) | -0.042**<br>(0.021) | -0.036<br>(0.023) | -0.033<br>(0.023) |
| Facility catchment population (1000) |  | 0.001***<br>(0.000) | 0.001***<br>(0.000) | 0.001***<br>(0.000) | 0.001***<br>(0.000) |
| Male healthcare worker |  |  | -0.040**<br>(0.016) | -0.044***<br>(0.016) | -0.043***<br>(0.016) |
| Medical Assistant |  |  | 0.023**<br>(0.012) | 0.025**<br>(0.011) | 0.025**<br>(0.011) |
| Nurse Officer/Midwife Technician |  |  | 0.065*<br>(0.034) | 0.070*<br>(0.037) | 0.067*<br>(0.037) |
| Professional experience (years) |  |  | -0.003***<br>(0.001) | -0.002***<br>(0.001) | -0.002***<br>(0.001) |
| Health condition: ALRI |  |  |  | -0.019**<br>(0.008) | -0.019**<br>(0.008) |
| Health condition: ANC |  |  |  | -0.041<br>(0.050) | -0.043<br>(0.050) |
| Health condition: Diarrhoea |  |  |  | 0.036<br>(0.022) | 0.036<br>(0.022) |
| Health condition: Maternal/Newborn healthcare |  |  |  | -0.033<br>(0.046) | -0.034<br>(0.046) |
| Health condition: NCDs |  |  |  | -0.018*<br>(0.010) | -0.019*<br>(0.010) |
| Health condition: Other |  |  |  | -0.030***<br>(0.008) | -0.031***<br>(0.008) |
| Patient order |  |  |  |  | -0.000**<br>(0.000) |
| Constant | 0.038*<br>(0.020) | -0.028<br>(0.021) | -0.055*<br>(0.030) | -0.057*<br>(0.029) | -0.050*<br>(0.030) |
| Observations | 3905 | 3905 | 3905 | 3905 | 3905 |
| R<sup>2</sup> | 0.036 | 0.068 | 0.080 | 0.086 | 0.087 |

*Notes:* This table presents the results from Equation 1, using data on consultations from healthcare-worker observations in outpatient departments in community hospitals and health centres participating in the Thanzi La Mawa (TLM) Time and Motion Study. The outcome variable is the likelihood of the healthcare worker being observed to conduct any type of diagnostics or monitoring activity during a patient consultation. FBP equals 1 if the observed consultation was conducted at a faith-based (CHAM) facility and 0 if conducted at a government facility. The omitted reference category for observation day is Observation Day 1, for facility type it is health centre, for healthcare-worker profession it is Clinical Officer/Technician; and for health condition it is malaria. ALRI = acute lower respiratory infection, ANC= antenatal care, NCD = non-communicable disease. Robust standard errors are reported in parentheses. \* $p < 0.10$ , \*\* $p < 0.05$ , \*\*\* $p < 0.01$ .

**Table 7:** Faith-based facility ownership and likelihood of healthcare workers dispensing/prescribing medication.

|  | Dependent variable: Dispensing/prescribing medication |  |  |  |  |
| --- | --- | --- | --- | --- | --- |
|  | (1) | (2) | (3) | (4) | (5) |
| FBP | 0.256***<br>(0.018) | 0.304***<br>(0.017) | 0.126***<br>(0.029) | 0.165***<br>(0.029) | 0.185***<br>(0.030) |
| Observation day 2 | 0.118***<br>(0.018) | 0.069***<br>(0.017) | 0.070***<br>(0.017) | 0.062***<br>(0.017) | 0.049***<br>(0.017) |
| Observation day 3+ | 0.118***<br>(0.018) | 0.075***<br>(0.018) | 0.085***<br>(0.018) | 0.078***<br>(0.018) | 0.079***<br>(0.017) |
| Observed during whole shift | 0.193***<br>(0.028) | 0.139***<br>(0.029) | 0.136***<br>(0.030) | 0.130***<br>(0.029) | 0.121***<br>(0.029) |
| Survey month: February | 0.043<br>(0.031) | 0.190***<br>(0.032) | -0.011<br>(0.043) | -0.004<br>(0.043) | 0.021<br>(0.043) |
| Survey month: March | -0.094***<br>(0.029) | 0.242***<br>(0.033) | -0.122**<br>(0.061) | -0.135**<br>(0.060) | -0.127**<br>(0.060) |
| Survey month: April | 0.075***<br>(0.029) | 0.413***<br>(0.034) | 0.033<br>(0.068) | 0.015<br>(0.068) | 0.041<br>(0.068) |
| Normal working day | 0.039**<br>(0.016) | 0.018<br>(0.015) | 0.017<br>(0.015) | 0.017<br>(0.015) | 0.024<br>(0.015) |
| Facility location: Rural |  | 0.198***<br>(0.016) | 0.118***<br>(0.028) | 0.127***<br>(0.028) | 0.145***<br>(0.028) |
| Facility type: Community Hospital |  | -0.257***<br>(0.025) | -0.146***<br>(0.034) | -0.113***<br>(0.034) | -0.129***<br>(0.035) |
| Facility catchment population (1000) |  | 0.003***<br>(0.000) | 0.002***<br>(0.000) | 0.002***<br>(0.000) | 0.002***<br>(0.000) |
| Male healthcare worker |  |  | -0.032<br>(0.026) | -0.044*<br>(0.026) | -0.053**<br>(0.026) |
| Medical Assistant |  |  | -0.197***<br>(0.025) | -0.187***<br>(0.025) | -0.190***<br>(0.025) |
| Nurse Officer/Midwife Technician |  |  | 0.232***<br>(0.046) | 0.295***<br>(0.046) | 0.316***<br>(0.046) |
| Professional experience (years) |  |  | 0.001<br>(0.002) | 0.003<br>(0.002) | 0.001<br>(0.002) |
| Health condition: ALRI |  |  |  | 0.023<br>(0.019) | 0.026<br>(0.019) |
| Health condition: ANC |  |  |  | -0.380***<br>(0.069) | -0.365***<br>(0.069) |
| Health condition: Diarrhoea |  |  |  | -0.057<br>(0.036) | -0.053<br>(0.036) |
| Health condition: Maternal/Newborn healthcare |  |  |  | -0.203***<br>(0.057) | -0.197***<br>(0.057) |
| Health condition: NCDs |  |  |  | -0.088***<br>(0.024) | -0.080***<br>(0.024) |
| Health condition: Other |  |  |  | -0.079***<br>(0.019) | -0.074***<br>(0.019) |
| Patient order |  |  |  |  | 0.001***<br>(0.000) |
| Constant | -0.013<br>(0.043) | -0.261***<br>(0.045) | 0.215***<br>(0.070) | 0.202***<br>(0.069) | 0.157**<br>(0.070) |
| Observations | 3905 | 3905 | 3905 | 3905 | 3905 |
| R<sup>2</sup> | 0.082 | 0.186 | 0.210 | 0.224 | 0.229 |

*Notes:* This table presents the results from Equation 1, using data on consultations from healthcare-worker observations in outpatient departments in community hospitals and health centres participating in the Thanzi La Mawa (TLM) Time and Motion Study. The outcome variable is the likelihood of the healthcare worker being observed to dispense or prescribe any medication during a patient consultation. FBP equals 1 if the observed consultation was conducted at a faith-based (CHAM) facility and 0 if conducted at a government facility. The omitted reference category for observation day is Observation Day 1, for facility type it is health centre, for healthcare-worker profession it is Clinical Officer/Technician; and for health condition it is malaria. ALRI = acute lower respiratory infection, ANC= antenatal care, NCD = non-communicable disease. Robust standard errors are reported in parentheses. \* $p < 0.10$ , \*\* $p < 0.05$ , \*\*\* $p < 0.01$ .

Our conclusions remain unchanged when restricting the analytical sample to observation day 2 or later, see Table A6 in the Appendix. The results are qualitatively similar to the main estimates when using wild cluster bootstrap standard errors at the facility level. We continue to observe statistically significant effects for physical examination and the coefficients for conducting diagnostics/monitoring and dispensing/prescribing drugs remain comparable in magnitude and direction. However, these latter estimates become marginally statistically insignificant (with p-values of 0.10 and 0.14, respectively). Moreover, our results on both consultation length and consultation contents hold after accounting for the number of patients seen per day, keeping in mind the endogenous nature of this variable, see Table A7 in the Appendix. The estimate on consultation time reduces to 0.96 minutes after adding this covariate but remains statistically significant. Our results are in line with Das et al. (2012) who report that high patient loads in public facilities in India can lead to rushed encounters.

### 5.2 Patient Reported Outcomes

Table 8 presents the regression results for patient reported primary care consultation contents when exiting the facility. The patient level analyses confirm that outpatient visits to CHAM facilities are more likely to result in a physical examination and a test. We also note that the coefficient on FBP remains relatively stable after adding patient-level covariates in columns 3 and 6, which suggests that case-mix variation is unlikely to explain our previously observed findings using direct healthcare worker observations. Moreover, clients interacting with FBPs are more likely to have been diagnosed with a new condition potentially reflecting the previously observed higher likelihood of an observed consultation consisting of any diagnostics or monitoring activity analysing direct healthcare worker observations. On the other hand, we do not observe a difference in patients reporting having received any treatment or medication, potentially explained by the broad interpretation of the question during interview and that the vast majority of all respondents reported to have received a treatment or medication. The results for patients reporting receiving a test and being diagnosed with a new condition remain consistent when applying wild cluster bootstrap standard errors at the facility level, see Table A8 in the Appendix. Although the coefficient on physical examination is positive, it does not reach statistical significance after correcting the standard errors.

**Table 8:** Care received during outpatient consultation reported by patients.

|  | Physical examination |  |  | Any test |  |  | Diagnosed with new condition |  |  | Any treatment or medication |  |  |
| --- | --- | --- | --- | --- | --- | --- | --- | --- | --- | --- | --- | --- |
|  | (1) | (2) | (3) | (4) | (5) | (6) | (7) | (8) | (9) | (10) | (11) | (12) |
| FBP | 0.164***<br>(0.032) | 0.087**<br>(0.034) | 0.083**<br>(0.039) | 0.105***<br>(0.032) | 0.130***<br>(0.035) | 0.135***<br>(0.034) | 0.084***<br>(0.019) | 0.109***<br>(0.017) | 0.140***<br>(0.023) | -0.001<br>(0.020) | -0.042**<br>(0.019) | -0.031<br>(0.022) |
| Facility location: Rural |  | -0.171***<br>(0.036) | -0.229***<br>(0.041) |  | -0.027<br>(0.037) | -0.051<br>(0.038) |  | 0.094***<br>(0.026) | 0.034<br>(0.030) |  | 0.058***<br>(0.021) | 0.048*<br>(0.027) |
| Facility type: Community Hospital |  | 0.138***<br>(0.039) | 0.074*<br>(0.042) |  | -0.064*<br>(0.039) | -0.052<br>(0.037) |  | 0.074***<br>(0.027) | 0.018<br>(0.031) |  | 0.013<br>(0.020) | 0.013<br>(0.024) |
| Facility catchment population (1000) |  | -0.001***<br>(0.000) | -0.002***<br>(0.000) |  | 0.000<br>(0.000) | 0.000<br>(0.000) |  | 0.002***<br>(0.000) | 0.002***<br>(0.000) |  | -0.001***<br>(0.000) | -0.001**<br>(0.000) |
| Patient age: 6-12 |  |  | -0.101*<br>(0.052) |  |  | -0.012<br>(0.044) |  |  | 0.030<br>(0.035) |  |  | 0.016<br>(0.033) |
| Patient age: 13-17 |  |  | -0.089<br>(0.072) |  |  | -0.018<br>(0.058) |  |  | 0.021<br>(0.045) |  |  | 0.074**<br>(0.031) |
| Patient age: 18-25 |  |  | -0.039<br>(0.063) |  |  | 0.053<br>(0.056) |  |  | 0.039<br>(0.045) |  |  | 0.027<br>(0.037) |
| Patient age: 26-49 |  |  | -0.098*<br>(0.059) |  |  | -0.018<br>(0.052) |  |  | 0.067<br>(0.042) |  |  | 0.044<br>(0.035) |
| Patient age: 50+ |  |  | -0.088<br>(0.065) |  |  | 0.023<br>(0.061) |  |  | 0.147***<br>(0.054) |  |  | 0.075*<br>(0.039) |
| Male Patient |  |  | -0.042<br>(0.029) |  |  | -0.042<br>(0.026) |  |  | 0.001<br>(0.020) |  |  | 0.018<br>(0.018) |
| Sought care for self |  |  | -0.025<br>(0.050) |  |  | -0.086**<br>(0.043) |  |  | 0.033<br>(0.037) |  |  | -0.004<br>(0.028) |
| Household wealth score |  |  | 0.012*<br>(0.007) |  |  | 0.009<br>(0.007) |  |  | 0.007<br>(0.005) |  |  | 0.003<br>(0.005) |
| Health condition: ALRI |  |  | -0.070<br>(0.045) |  |  | -0.469***<br>(0.037) |  |  | -0.009<br>(0.028) |  |  | -0.117***<br>(0.028) |
| Health condition: Diarrhoea |  |  | -0.256***<br>(0.059) |  |  | -0.464***<br>(0.061) |  |  | -0.022<br>(0.046) |  |  | -0.083**<br>(0.039) |
| Health condition: NCDs |  |  | 0.078<br>(0.058) |  |  | -0.506***<br>(0.053) |  |  | 0.062<br>(0.049) |  |  | -0.130***<br>(0.035) |
| Health condition: Other |  |  | -0.017<br>(0.036) |  |  | -0.445***<br>(0.026) |  |  | -0.029<br>(0.023) |  |  | -0.107***<br>(0.019) |
| Health status: Mild symptoms |  |  | 0.429***<br>(0.085) |  |  | 0.466***<br>(0.071) |  |  | 0.051<br>(0.036) |  |  | 0.032<br>(0.063) |
| Health status: Moderate symptoms |  |  | 0.468***<br>(0.089) |  |  | 0.419***<br>(0.076) |  |  | 0.077*<br>(0.041) |  |  | 0.034<br>(0.066) |
| Health status: Severe/Life-threatening symptoms |  |  | 0.475***<br>(0.108) |  |  | 0.428***<br>(0.099) |  |  | 0.185***<br>(0.067) |  |  | -0.017<br>(0.081) |
| Constant | 0.392***<br>(0.026) | 0.504***<br>(0.051) | 0.280***<br>(0.088) | 0.579***<br>(0.027) | 0.609***<br>(0.053) | 0.570***<br>(0.073) | 0.067***<br>(0.014) | -0.102***<br>(0.030) | -0.166***<br>(0.041) | 0.898***<br>(0.016) | 0.927***<br>(0.025) | 0.936***<br>(0.063) |
| Observations | 1118 | 1118 | 1118 | 1118 | 1118 | 1118 | 1118 | 1118 | 1118 | 1118 | 1118 | 1118 |
| R<sup>2</sup> | 0.023 | 0.098 | 0.148 | 0.010 | 0.014 | 0.244 | 0.014 | 0.046 | 0.094 | 0.000 | 0.026 | 0.061 |

*Notes:* This table presents the results from Equation 1, using data from the patient-exit interviews conducted as part of the Thanzi La Mawa (TLM) project with patients leaving community hospitals and health centres who reported attending an outpatient department. The outcome variables capture the likelihood that a patient reported receiving a physical examination, any diagnostic test, a new diagnosis, or any treatment/medication during their visit. FBP equals 1 if the patient attended a consultation at a faith-based (CHAM) facility and 0 if they visited a government facility. The omitted reference category for facility type is health centre, for patient age is 0–5 years, and for health condition is malaria. ALRI = acute lower respiratory infection; NCD = non-communicable disease. Robust standard errors in parentheses. \* $p < .10$ , \*\* $p < .05$ , \*\*\* $p < .01$ .

For the reduced sample of patients successfully contacted for a telephone follow-up, respondents who visited a FBP are more likely to report that their treatment is working compared with those who visited a government provider, see columns 4–6 in Table 9). Although the coefficient on faith-based facility ownership is positive for the likelihood of patients reporting that their sickness has resolved, it does not reach conventional levels of statistical significance, see columns 1–3 in Table 9.

**Table 9:**
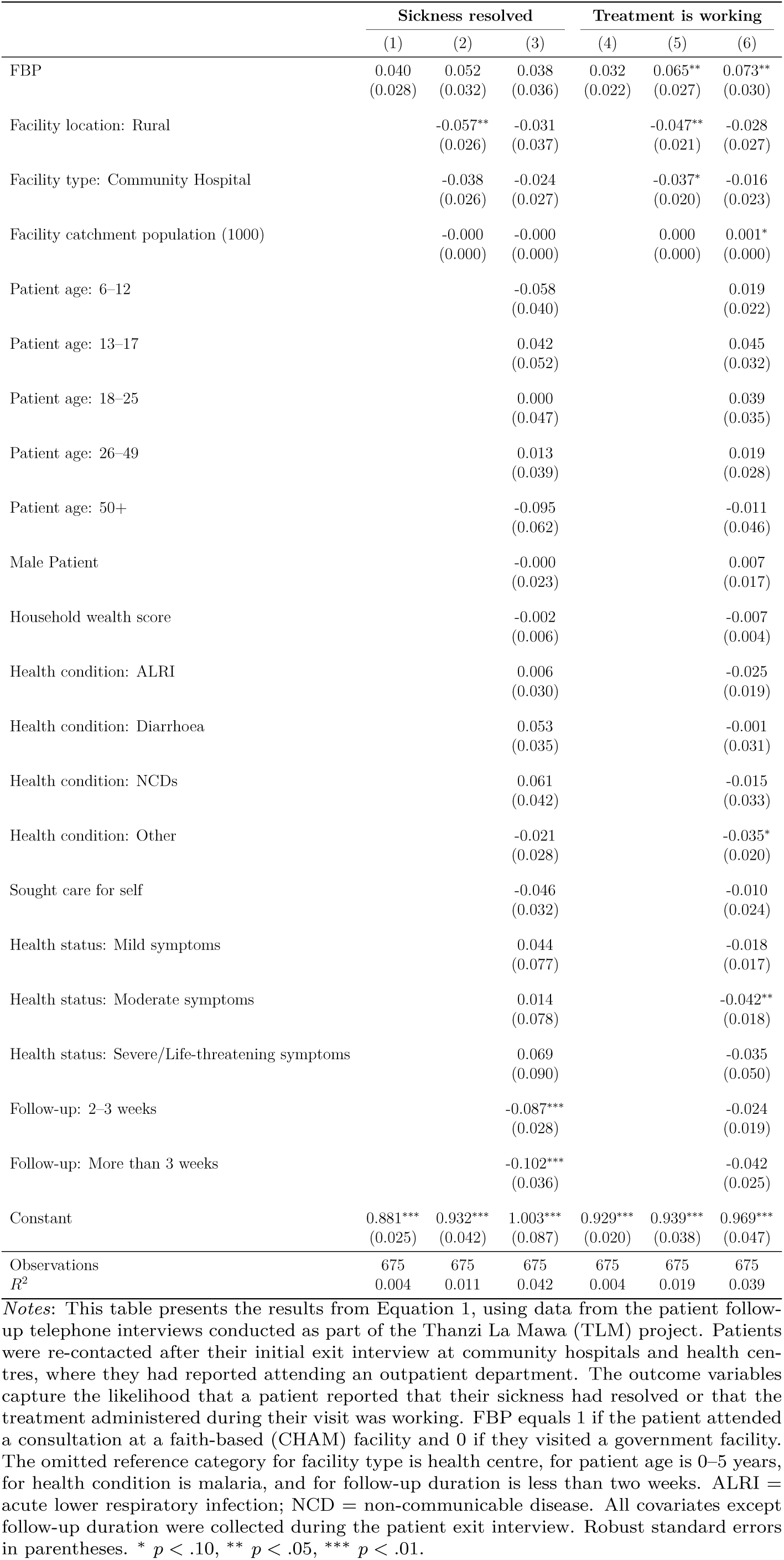
Patient reported outcomes at follow-up.

|  | Sickness resolved |  |  | Treatment is working |  |  |
| --- | --- | --- | --- | --- | --- | --- |
|  | (1) | (2) | (3) | (4) | (5) | (6) |
| FBP | 0.040<br>(0.028) | 0.052<br>(0.032) | 0.038<br>(0.036) | 0.032<br>(0.022) | 0.065**<br>(0.027) | 0.073**<br>(0.030) |
| Facility location: Rural |  | -0.057**<br>(0.026) | -0.031<br>(0.037) |  | -0.047**<br>(0.021) | -0.028<br>(0.027) |
| Facility type: Community Hospital |  | -0.038<br>(0.026) | -0.024<br>(0.027) |  | -0.037*<br>(0.020) | -0.016<br>(0.023) |
| Facility catchment population (1000) |  | -0.000<br>(0.000) | -0.000<br>(0.000) |  | 0.000<br>(0.000) | 0.001*<br>(0.000) |
| Patient age: 6–12 |  |  | -0.058<br>(0.040) |  |  | 0.019<br>(0.022) |
| Patient age: 13–17 |  |  | 0.042<br>(0.052) |  |  | 0.045<br>(0.032) |
| Patient age: 18–25 |  |  | 0.000<br>(0.047) |  |  | 0.039<br>(0.035) |
| Patient age: 26–49 |  |  | 0.013<br>(0.039) |  |  | 0.019<br>(0.028) |
| Patient age: 50+ |  |  | -0.095<br>(0.062) |  |  | -0.011<br>(0.046) |
| Male Patient |  |  | -0.000<br>(0.023) |  |  | 0.007<br>(0.017) |
| Household wealth score |  |  | -0.002<br>(0.006) |  |  | -0.007<br>(0.004) |
| Health condition: ALRI |  |  | 0.006<br>(0.030) |  |  | -0.025<br>(0.019) |
| Health condition: Diarrhoea |  |  | 0.053<br>(0.035) |  |  | -0.001<br>(0.031) |
| Health condition: NCDs |  |  | 0.061<br>(0.042) |  |  | -0.015<br>(0.033) |
| Health condition: Other |  |  | -0.021<br>(0.028) |  |  | -0.035*<br>(0.020) |
| Sought care for self |  |  | -0.046<br>(0.032) |  |  | -0.010<br>(0.024) |
| Health status: Mild symptoms |  |  | 0.044<br>(0.077) |  |  | -0.018<br>(0.017) |
| Health status: Moderate symptoms |  |  | 0.014<br>(0.078) |  |  | -0.042**<br>(0.018) |
| Health status: Severe/Life-threatening symptoms |  |  | 0.069<br>(0.090) |  |  | -0.035<br>(0.050) |
| Follow-up: 2–3 weeks |  |  | -0.087***<br>(0.028) |  |  | -0.024<br>(0.019) |
| Follow-up: More than 3 weeks |  |  | -0.102***<br>(0.036) |  |  | -0.042<br>(0.025) |
| Constant | 0.881***<br>(0.025) | 0.932***<br>(0.042) | 1.003***<br>(0.087) | 0.929***<br>(0.020) | 0.939***<br>(0.038) | 0.969***<br>(0.047) |
| Observations | 675 | 675 | 675 | 675 | 675 | 675 |
| $R^2$ | 0.004 | 0.011 | 0.042 | 0.004 | 0.019 | 0.039 |

*Notes:* This table presents the results from Equation 1, using data from the patient follow-up telephone interviews conducted as part of the Thanzi La Mawa (TLM) project. Patients were re-contacted after their initial exit interview at community hospitals and health centres, where they had reported attending an outpatient department. The outcome variables capture the likelihood that a patient reported that their sickness had resolved or that the treatment administered during their visit was working. FBP equals 1 if the patient attended a consultation at a faith-based (CHAM) facility and 0 if they visited a government facility. The omitted reference category for facility type is health centre, for patient age is 0–5 years, for health condition is malaria, and for follow-up duration is less than two weeks. ALRI = acute lower respiratory infection; NCD = non-communicable disease. All covariates except follow-up duration were collected during the patient exit interview. Robust standard errors in parentheses. \* $p < .10$ , \*\* $p < .05$ , \*\*\* $p < .01$ .

## 6 Discussion and Conclusion

This study provides new evidence on differences in the quality of primary healthcare between faith-based and government-managed facilities in Malawi. Using direct observations of healthcare worker behaviour through a time-and-motion methodology, complemented by patient-reported data, we document systematic differences in consultation time, processes and patient-reported outcomes across ownership types.

Relying on direct healthcare worker observations, we find that providers in faith-based facilities spend on average 1.9 additional minutes with each patient. This represents a sizeable effect given the mean consultation duration of 2.52 minutes. Faith-based facilities are also characterised by a higher likelihood of conducting physical examinations, undertaking diagnostic and monitoring activities, and prescribing or dispensing medication. These patterns are consistent with evidence from Tanzania based on direct clinician observations (Leonard et al., 2007), which indicates that private and NGO providers tend to deliver higher process quality, particularly in diagnostic effort and provider–patient communication.

Analyses based on patient-exit interviews reinforce these findings. Patients attending faith-based facilities are more likely to report receiving physical examinations, tests, or new diagnoses during their visit. Follow-up interviews also indicate higher self-reported treatment effectiveness among patients who sought care at faith-based providers. Taken together, these patterns suggest that process quality differences observed during clinical encounters may translate into better patient-perceived outcomes.

The mechanisms behind these ownership differences are consistent with prior literature linking longer consultations and more comprehensive assessments to improved diagnostic accuracy and patient outcomes (Leonard and Masatu, 2010; Das et al., 2016; Okeke, 2021). Okeke (2021), for instance, show that exogenous increases in consultation time lead to more detailed history taking, more physical examinations, increased diagnostic testing, and higher rates of correct diagnosis and treatment. Faith-based providers in this study may have greater capacity to deliver high-quality care because they operate further from their productivity frontiers (Duchoslav and Cecchi, 2019), consistent with the lower patient volumes observed in our data and documented elsewhere (Mæstad et al., 2010; Sheffel et al., 2024). While we account for patient order and caseload per provider, organisational differences in incentives and governance structures may also drive effort. Prior research suggests that faith-based providers often embed mission-driven, pro-social motivations within their organisational culture (Olivier et al., 2015; Reinikka and Svensson, 2010; Bjorvatn and Svensson, 2016; Bawuah et al., 2024). This may contribute to a smaller “know–do gap”, the discrepancy between provider competence and actual effort, which has been found to be narrower among non-governmental providers in similar settings (Leonard et al., 2007; Das et al., 2008; Leonard and Masatu, 2010). Although our study does not measure competence directly, evidence from Malawi using clinical vignettes finds no systematic differences in competence between faith-based and public providers (Tafesse and Chalkley, 2024). The differences observed here in consultation time and process quality are therefore more plausibly interpreted as variation in effort exerted during patient interactions.

At the same time, our results also highlight system-wide constraints. The average consultation duration of 2.5 minutes across all facilities is well below the WHO’s ten-minute recommendation and is shorter than reported in comparable LMIC settings (Irving et al., 2017; Okeke, 2021; Kovacs and Lagarde, 2022). Consistent with Das and Sohnesen (2006), consultation time declines over the course of the working day. We further show that higher daily caseloads per provider are associated with shorter consultations. Although we cannot establish causal effects, the results suggest that heavy workloads may undermine care quality in Malawi, contrary to evidence from other Sub-Saharan African countries (Mæstad et al., 2010; Kovacs and Lagarde, 2022; Daniels et al., 2022), where lower daily caseloads may mitigate these pressures.

This study has several limitations. Although the number of observed patient consultations is large, the analytical sample is restricted to 19 facilities. Analysing data on direct healthcare worker observations throughout multiple shifts allows us to capture real and complex patient interactions and patient flows during the day, representative for outpatient primary care provision in Malawi. Supplementary evidence from patient-exit interviews shows that our main conclusions are robust to controlling for a wide range of observable patient characteristics, including health condition and self-reported severity. However, as with all studies relying on direct observations and patient recall rather than standardised patients, we cannot infer causal effects due to potential case-mix differences (Peabody et al., 2000). Moreover, conducting more activities during consultations may not necessarily reflect appropriate care. Over-provision remains a concern in SSA, including in faith-based facilities (King et al., 2021, 2023). Finally, we are unable to link facility ownership to objectively measured health outcomes. However, recent evidence suggests improved health outcomes among patients attending faith-based tertiary facilities in the region (Parker et al., 2025).

Taken together, these findings contribute to a growing body of evidence on the role of non-state actors in LMIC health systems. Faith-based facilities, which account for a substantial share of Malawi’s healthcare infrastructure and play an important role in rural and underserved areas, appear to complement public provision by offering more comprehensive primary care. Quality differences by ownership, however, are likely to vary across service areas. For instance, faith-based facilities in Malawi have been shown to be less likely to adhere to sexual and reproductive health guidelines (Tafesse and Chalkley, 2021). The consistently short consultation durations observed across all facilities point to systemic constraints that cut across ownership categories. Strengthening primary care in Malawi will therefore require sustained investment in human resources, improved workload management, and the alignment of financial incentives with quality improvement goals in both public and faith-based sectors.

## Data Availability

Anonymised and non-sensitive data produced in the present study are available upon reasonable request to the authors.

## A Appendix

**Table A1:** Descriptive Statistics: Shift-level observations by facility ownership.

|  | **Faith-based** |  | **Government** |  |
| --- | --- | --- | --- | --- |
|  | *Mean* | *SD* | *Mean* | *SD* |
| Day of observation | 1.86 | 0.83 | 1.91 | 0.84 |
| Observed during whole shift | 0.88 | 0.33 | 0.94 | 0.24 |
| Normal working day | 0.60 | 0.49 | 0.42 | 0.50 |
| Survey month: January | 0.10 | 0.30 | 0.06 | 0.24 |
| Survey month: February | 0.26 | 0.44 | 0.18 | 0.39 |
| Survey month: March | 0.40 | 0.49 | 0.21 | 0.42 |
| Survey month: April | 0.24 | 0.43 | 0.55 | 0.51 |
| Facility location: Rural | 0.60 | 0.49 | 0.58 | 0.50 |
| Facility type: Community Hospital | 0.58 | 0.50 | 0.64 | 0.49 |
| Facility type: Health Centre | 0.42 | 0.50 | 0.36 | 0.49 |
| Facility catchment population (1000) | 26.67 | 11.86 | 55.91 | 44.41 |
| Number of patients seen/shift | 27.26 | 17.49 | 80.70 | 53.84 |
| Clinical Officer / Technician | 0.50 | 0.51 | 0.42 | 0.50 |
| Medical Assistant | 0.16 | 0.37 | 0.52 | 0.51 |
| Nurse Midwife Technician | 0.18 | 0.39 | 0.03 | 0.17 |
| Nurse Officer | 0.16 | 0.37 | 0.03 | 0.17 |
| Male healthcare worker | 0.58 | 0.50 | 0.58 | 0.50 |
| Professional experience (years) | 6.40 | 5.92 | 6.91 | 6.09 |
| Observations | 50 |  | 33 |  |

*Notes:* This table reports descriptive statistics by facility ownership at the healthcare-worker-shift level for the analytical sample of observed outpatient consultations conducted in outpatient departments of community hospitals and health centres included in the Thanzi La Mawa (TLM) project Time and Motion Study (TMS). Mean/proportion and standard deviation (SD) values are presented.

**Table A2:**
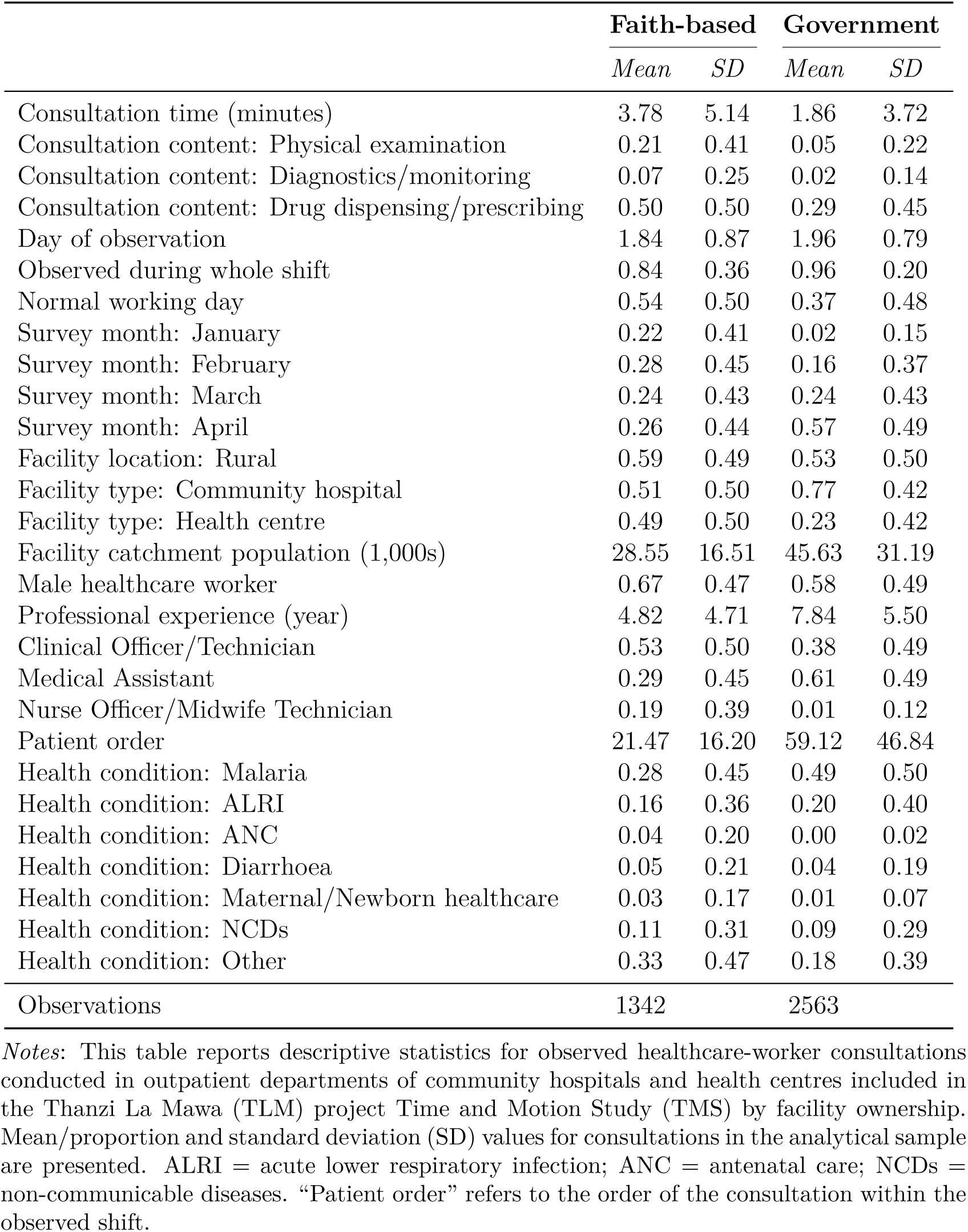
Descriptive statistics by facility ownership: Consultation observations.

**Table A3:** Descriptive statistics by facility ownership: Patient exit interview.

|  | Faith-based provider (FBP) |  | Public provider |  |
| --- | --- | --- | --- | --- |
|  | Mean | SD | Mean | SD |
| Patient reported receiving: Physical examination | 0.56 | 0.50 | 0.39 | 0.49 |
| Patient reported receiving: Any test | 0.68 | 0.47 | 0.58 | 0.49 |
| Patient reported receiving: New diagnosis | 0.15 | 0.36 | 0.07 | 0.25 |
| Patient reported receiving: Any treatment/medication | 0.90 | 0.30 | 0.90 | 0.30 |
| Facility location: Rural | 0.53 | 0.50 | 0.45 | 0.50 |
| Facility type: Community Hospital | 0.64 | 0.48 | 0.30 | 0.46 |
| Patient age: 0–5 | 0.43 | 0.49 | 0.21 | 0.41 |
| Patient age: 6–12 | 0.11 | 0.31 | 0.07 | 0.26 |
| Patient age: 13–17 | 0.05 | 0.22 | 0.10 | 0.30 |
| Patient age: 18–25 | 0.11 | 0.31 | 0.21 | 0.41 |
| Patient age: 26–49 | 0.21 | 0.41 | 0.32 | 0.47 |
| Patient age: 50+ | 0.09 | 0.29 | 0.10 | 0.30 |
| Male Patient | 0.46 | 0.50 | 0.38 | 0.49 |
| Patient sought care for self | 0.41 | 0.49 | 0.69 | 0.46 |
| Household wealth quintile: Poorest | 0.27 | 0.45 | 0.40 | 0.49 |
| Household wealth quintile: Poor | 0.15 | 0.36 | 0.22 | 0.41 |
| Household wealth quintile: Middle | 0.18 | 0.38 | 0.18 | 0.39 |
| Household wealth quintile: Richer | 0.17 | 0.38 | 0.14 | 0.35 |
| Household wealth quintile: Richest | 0.22 | 0.41 | 0.05 | 0.22 |
| Health condition: Malaria | 0.27 | 0.44 | 0.27 | 0.44 |
| Health condition: ALRI | 0.18 | 0.38 | 0.18 | 0.39 |
| Health condition: Diarrhoea | 0.06 | 0.24 | 0.04 | 0.21 |
| Health condition: NCDs | 0.10 | 0.30 | 0.07 | 0.26 |
| Health condition: Other | 0.39 | 0.49 | 0.43 | 0.50 |
| Health status: Healthy | 0.06 | 0.23 | 0.01 | 0.08 |
| Health status: Mild symptoms | 0.50 | 0.50 | 0.56 | 0.50 |
| Health status: Moderate symptoms | 0.40 | 0.49 | 0.36 | 0.48 |
| Health status: Severe/Life-threatening symptoms | 0.05 | 0.21 | 0.07 | 0.26 |
| Observations | 776 |  | 342 |  |

*Notes:* This table reports descriptive statistics for adults sampled for a patient-exit interview as part of the Thanzi La Mawa (TLM) project data collection. Respondents were interviewed as they left community hospitals and health centres and reported attending an outpatient department. Statistics are presented by facility ownership. Mean/proportion and standard deviation (SD) values are reported for patients in the analytical sample. FBP is a binary indicator equal to 1 if the patient attended a faith-based (CHAM) facility and 0 if they attended a government facility. “Patient sought care for self” equals 1 if the respondent sought care for themselves and 0 if they accompanied someone seeking care. Wealth quintiles are constructed using household asset ownership. Illness severity categories reflect patients’ self-reported symptoms at the time of exit. ALRI = acute lower respiratory infection; NCD = non-communicable disease.

**Table A4:**
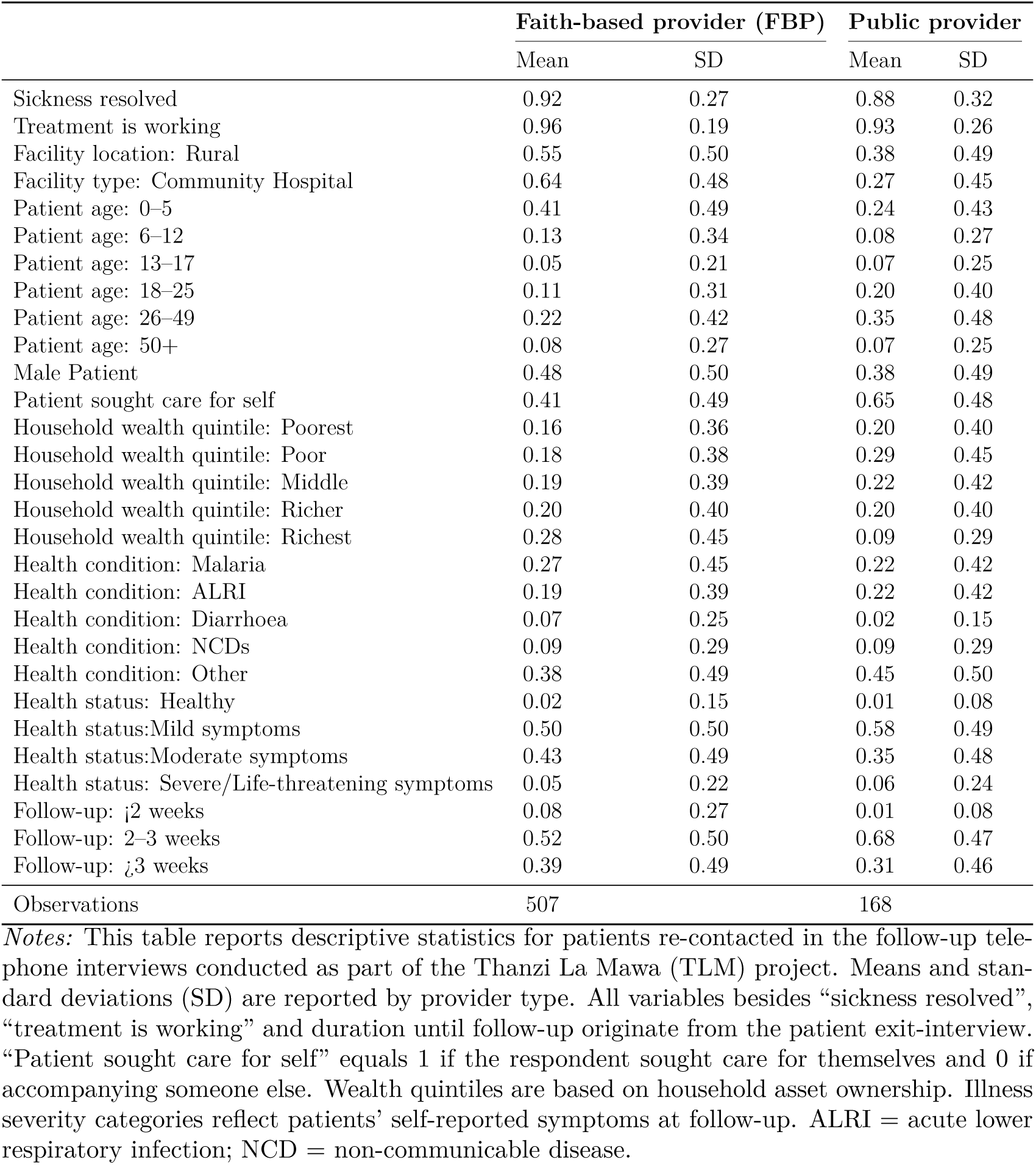
Descriptive statistics by facility ownership: Patient follow-up.

**Table A5:** Robustness: Faith-based facility ownership and consultation time.

|  | Whole shift observation |  | < 30 min consultation |  | > 1 day observation |  | Wild cluster bootstrap |  |
| --- | --- | --- | --- | --- | --- | --- | --- | --- |
|  | (1)<br>Consultation time | (2)<br>Consultation time | (3)<br>Consultation time | (4)<br>Consultation time | (5)<br>Consultation time | (6)<br>Consultation time | (7)<br>Consultation time | (8)<br>Consultation time |
| FBP | 2.218***<br>(0.400) | 1.960***<br>(0.395) | 2.015***<br>(0.304) | 1.775***<br>(0.301) | 2.445***<br>(0.575) | 2.319***<br>(0.565) | 2.189**<br>(1.023) | 1.907*<br>(0.920) |
| Observation day 2 | -0.115<br>(0.185) | 0.068<br>(0.189) | -0.170<br>(0.139) | -0.020<br>(0.140) |  |  | -0.258<br>(0.268) | -0.081<br>(0.229) |
| Observation day 3+ | -0.124<br>(0.178) | -0.151<br>(0.177) | -0.137<br>(0.136) | -0.144<br>(0.135) | 0.005<br>(0.171) | -0.162<br>(0.180) | -0.178<br>(0.338) | -0.186<br>(0.331) |
| Observed during whole shift |  |  | 0.315<br>(0.229) | 0.422*<br>(0.228) | 1.374***<br>(0.329) | 1.476***<br>(0.328) | 0.340<br>(0.400) | 0.465<br>(0.368) |
| Survey month: February | 2.579***<br>(0.505) | 2.252***<br>(0.502) | 2.092***<br>(0.360) | 1.791***<br>(0.359) | 2.712***<br>(0.520) | 2.453***<br>(0.512) | 2.289**<br>(1.058) | 1.937*<br>(0.954) |
| Survey month: March | 1.290<br>(0.789) | 1.202<br>(0.784) | 1.009*<br>(0.546) | 0.907*<br>(0.543) | 0.612<br>(1.123) | 0.597<br>(1.116) | 1.098<br>(1.395) | 0.978<br>(1.218) |
| Survey month: April | 0.961<br>(0.845) | 0.602<br>(0.842) | 0.616<br>(0.596) | 0.298<br>(0.598) | 0.259<br>(1.240) | 0.062<br>(1.230) | 0.740<br>(1.342) | 0.367<br>(1.263) |
| Normal working day | 0.052<br>(0.156) | -0.066<br>(0.154) | 0.129<br>(0.112) | 0.044<br>(0.111) | 0.527*<br>(0.274) | 0.288<br>(0.283) | 0.120<br>(0.164) | 0.019<br>(0.143) |
| Facility location: Rural | 0.888***<br>(0.341) | 0.660*<br>(0.342) | 0.572**<br>(0.262) | 0.358<br>(0.263) | 0.423<br>(0.404) | 0.286<br>(0.405) | 0.630<br>(0.520) | 0.379<br>(0.484) |
| Facility type: Community Hospital | 1.696***<br>(0.519) | 1.913***<br>(0.519) | 1.617***<br>(0.383) | 1.804***<br>(0.386) | 1.963***<br>(0.703) | 2.194***<br>(0.699) | 1.778**<br>(0.742) | 1.999**<br>(0.704) |
| Facility catchment population (1000) | 0.028***<br>(0.005) | 0.027***<br>(0.005) | 0.025***<br>(0.004) | 0.024***<br>(0.004) | 0.014**<br>(0.006) | 0.016***<br>(0.006) | 0.028**<br>(0.012) | 0.027**<br>(0.010) |
| Male healthcare worker | -0.646*<br>(0.338) | -0.510<br>(0.338) | -0.171<br>(0.249) | -0.060<br>(0.249) | -0.973*<br>(0.504) | -0.793<br>(0.506) | -0.443<br>(0.440) | -0.312<br>(0.401) |
| Medical Assistant | -0.033<br>(0.314) | 0.054<br>(0.312) | 0.018<br>(0.219) | 0.046<br>(0.218) | -0.356<br>(0.588) | -0.220<br>(0.592) | -0.033<br>(0.694) | -0.000<br>(0.626) |
| Nurse Officer/Midwife Technician | 2.147***<br>(0.756) | 1.869**<br>(0.753) | 2.103***<br>(0.614) | 1.861***<br>(0.609) | 1.938**<br>(0.851) | 1.787**<br>(0.851) | 2.139**<br>(0.976) | 1.854**<br>(0.852) |
| Professional experience (years) | -0.024<br>(0.020) | 0.004<br>(0.020) | -0.018<br>(0.016) | 0.002<br>(0.017) | 0.010<br>(0.025) | 0.028<br>(0.026) | -0.012<br>(0.060) | 0.012<br>(0.053) |
| Health condition: ALRI | 0.511***<br>(0.155) | 0.484***<br>(0.154) | 0.611***<br>(0.122) | 0.582***<br>(0.120) | 0.569***<br>(0.156) | 0.537***<br>(0.155) | 0.555**<br>(0.204) | 0.520**<br>(0.191) |
| Health condition: ANC | -2.502***<br>(0.711) | -2.740***<br>(0.718) | -1.902***<br>(0.643) | -2.083***<br>(0.646) | -2.666**<br>(1.128) | -2.849**<br>(1.135) | -2.323<br>(1.378) | -2.535*<br>(1.380) |
| Health condition: Diarrhoea | 1.210***<br>(0.455) | 1.160***<br>(0.450) | 0.887***<br>(0.289) | 0.845***<br>(0.286) | 1.543**<br>(0.716) | 1.457**<br>(0.705) | 1.095*<br>(0.538) | 1.042*<br>(0.504) |
| Health condition: Maternal/Newborn healthcare | 1.041<br>(0.872) | 0.959<br>(0.857) | 1.272<br>(0.824) | 1.197<br>(0.813) | 3.835**<br>(1.497) | 3.564**<br>(1.500) | 1.034<br>(1.565) | 0.945<br>(1.465) |
| Health condition: NCDs | 2.614***<br>(0.346) | 2.498***<br>(0.343) | 2.200***<br>(0.253) | 2.107***<br>(0.250) | 3.022***<br>(0.411) | 2.930***<br>(0.410) | 2.503***<br>(0.331) | 2.391***<br>(0.320) |
| Health condition: Other | 1.690***<br>(0.207) | 1.612***<br>(0.204) | 1.586***<br>(0.149) | 1.526***<br>(0.148) | 1.778***<br>(0.226) | 1.735***<br>(0.223) | 1.667***<br>(0.272) | 1.596***<br>(0.253) |
| Patient order |  | -0.012***<br>(0.001) |  | -0.011***<br>(0.001) |  | -0.010***<br>(0.002) |  | -0.012***<br>(0.004) |
| Constant | -2.565***<br>(0.881) | -1.905**<br>(0.868) | -2.583***<br>(0.642) | -2.035***<br>(0.633) | -3.103**<br>(1.315) | -2.781**<br>(1.292) | -2.761<br>(1.905) | -2.116<br>(1.730) |
| Observations | 3582 | 3582 | 3893 | 3893 | 2410 | 2410 | 3905 | 3905 |
| R<sup>2</sup> | 0.137 | 0.147 | 0.179 | 0.191 | 0.180 | 0.189 |  |  |

*Notes:* This table presents the results from Equation 1, using data on consultations from healthcare-worker observations in outpatient departments in community hospitals and health centres participating in the Thanzi La Mawa (TLM) Time and Motion Study. The outcome variable is consultation time (minutes). FBP equals 1 if the observed consultation was conducted at a faith-based (CHAM) facility and 0 if conducted at a government facility. The omitted reference category for observation day is Observation Day 1, for facility type it is health centre, for healthcare-worker profession it is Clinical Officer/Technician; and for health condition it is malaria. ALRI = acute lower respiratory infection, ANC= antenatal care, NCD = non-communicable disease. Robust standard errors are reported in parentheses. Columns (1)–(2) restrict the analytical sample to healthcare workers who were observed during an entire shift. Columns (3)–(4) provide estimation results for consultations under 30 minutes. Columns (5)–(6) provide regression results for consultations observed during day 2-5 of observation. Columns (7)–(8) report wild cluster bootstrap standard errors at the facility level (999 replications) for the coefficient on FBP. Columns (1)–(6) report heteroskedasticity-robust standard errors. Standard errors in parentheses. \* $p < .10$ , \*\* $p < .05$ , \*\*\* $p < .01$ .

**Table A6:** Robustness: Faith-based facility ownership and observed consultation activities.

|  | > 1 day observation |  |  | Wild cluster bootstrap |  |  |
| --- | --- | --- | --- | --- | --- | --- |
|  | (1)<br>Physical exam | (2)<br>Diagnostics | (3)<br>Drug prescribing | (4)<br>Physical exam | (5)<br>Diagnostics | (6)<br>Drug prescribing |
| FBP | 0.256***<br>(0.036) | 0.137***<br>(0.022) | 0.296***<br>(0.050) | 0.132**<br>(0.046) | 0.073<br>(0.042) | 0.185<br>(0.120) |
| Observation day 3+ | -0.049***<br>(0.012) | -0.002<br>(0.005) | 0.040**<br>(0.019) | -0.014<br>(0.017) | -0.029<br>(0.017) | 0.079*<br>(0.039) |
| Observed during whole shift | 0.077***<br>(0.025) | 0.013<br>(0.014) | 0.069*<br>(0.039) | 0.002<br>(0.040) | 0.020<br>(0.020) | 0.121<br>(0.090) |
| Survey month: February | -0.002<br>(0.045) | 0.091***<br>(0.023) | -0.020<br>(0.058) | -0.011<br>(0.057) | 0.051<br>(0.042) | 0.021<br>(0.164) |
| Survey month: March | 0.194***<br>(0.074) | 0.235***<br>(0.041) | 0.061<br>(0.093) | 0.035<br>(0.083) | 0.059<br>(0.074) | -0.127<br>(0.254) |
| Survey month: April | 0.055<br>(0.076) | 0.270***<br>(0.043) | 0.202*<br>(0.107) | -0.084<br>(0.098) | 0.068<br>(0.088) | 0.041<br>(0.266) |
| Normal working day | -0.042**<br>(0.019) | -0.011<br>(0.011) | 0.016<br>(0.027) | 0.017<br>(0.019) | -0.030*<br>(0.017) | 0.024<br>(0.029) |
| Facility location: Rural | 0.045<br>(0.029) | 0.091***<br>(0.017) | 0.187***<br>(0.042) | 0.032<br>(0.043) | 0.051<br>(0.036) | 0.145*<br>(0.077) |
| Facility type: Community Hospital | 0.010<br>(0.043) | -0.174***<br>(0.032) | -0.275***<br>(0.052) | 0.073*<br>(0.039) | -0.033<br>(0.042) | -0.129<br>(0.130) |
| Facility catchment population (1000) | 0.003***<br>(0.000) | 0.001***<br>(0.000) | 0.004***<br>(0.001) | 0.001***<br>(0.000) | 0.001***<br>(0.000) | 0.002*<br>(0.001) |
| Male healthcare worker | 0.101***<br>(0.027) | -0.104***<br>(0.021) | -0.046<br>(0.040) | 0.031<br>(0.045) | -0.043<br>(0.044) | -0.053<br>(0.058) |
| Medical Assistant | 0.136***<br>(0.031) | 0.044***<br>(0.016) | -0.072<br>(0.046) | 0.011<br>(0.027) | 0.025<br>(0.024) | -0.190*<br>(0.098) |
| Nurse Officer/Midwife Technician | -0.025<br>(0.059) | -0.150***<br>(0.036) | 0.217***<br>(0.064) | 0.025<br>(0.061) | 0.067<br>(0.084) | 0.316*<br>(0.168) |
| Professional experience (years) | 0.007***<br>(0.002) | -0.000<br>(0.001) | 0.008***<br>(0.003) | 0.005<br>(0.003) | -0.002<br>(0.002) | 0.001<br>(0.004) |
| Health condition: ALRI | 0.068***<br>(0.014) | -0.009<br>(0.007) | 0.072***<br>(0.024) | 0.062***<br>(0.019) | -0.019*<br>(0.011) | 0.026<br>(0.173) |
| Health condition: ANC | 0.452***<br>(0.094) | 0.077<br>(0.073) | -0.257**<br>(0.113) | 0.456**<br>(0.163) | -0.043<br>(0.090) | -0.365***<br>(0.102) |
| Health condition: Diarrhoea | 0.095***<br>(0.036) | 0.085***<br>(0.032) | -0.046<br>(0.050) | 0.066**<br>(0.028) | 0.036<br>(0.053) | -0.053<br>(0.122) |
| Health condition: Maternal/Newborn healthcare | 0.133<br>(0.091) | 0.074<br>(0.072) | -0.316***<br>(0.094) | 0.038<br>(0.083) | -0.034<br>(0.075) | -0.197<br>(0.135) |
| Health condition: NCDs | 0.091***<br>(0.023) | -0.001<br>(0.011) | -0.037<br>(0.030) | 0.081***<br>(0.023) | -0.019<br>(0.016) | -0.080<br>(0.147) |
| Health condition: Other | 0.085***<br>(0.016) | -0.012*<br>(0.007) | -0.028<br>(0.024) | 0.093***<br>(0.019) | -0.031**<br>(0.014) | -0.074<br>(0.108) |
| Patient order | -0.001***<br>(0.000) | -0.000<br>(0.000) | 0.001***<br>(0.000) | -0.001***<br>(0.000) | -0.000<br>(0.000) | 0.001**<br>(0.000) |
| Observation day 2 |  |  |  | 0.016<br>(0.029) | -0.019<br>(0.016) | 0.049<br>(0.034) |
| Constant | -0.414***<br>(0.088) | -0.147***<br>(0.034) | -0.042<br>(0.120) | -0.112<br>(0.093) | -0.050<br>(0.065) | 0.157<br>(0.292) |
| Observations | 2410 | 2410 | 2410 | 3905 | 3905 | 3905 |
| $R^2$ | 0.194 | 0.134 | 0.282 | | | |

*Notes:* This table presents the results from Equation 1, using data on consultations from healthcare-worker observations in outpatient departments in community hospitals and health centres participating in the Thanzi La Mawa (TLM) Time and Motion Study. The outcome variables are the content of the observed consultation. FBP equals 1 if the observed consultation was conducted at a faith-based (CHAM) facility and 0 if conducted at a government facility. The omitted reference category for observation day is Observation Day 1, for facility type it is health centre, for healthcare-worker profession it is Clinical Officer/Technician; and for health condition it is malaria. ALRI = acute lower respiratory infection, ANC = antenatal care, NCD = non-communicable disease. Columns (1)–(3) restrict the sample to healthcare workers observed for more than one day and report heteroskedasticity-robust standard errors. Columns (4)–(6) report wild cluster bootstrap (999 replications) standard errors for the coefficient on FBP, and significance stars in these columns are based on the bootstrap $p$ -values. \* $p < .10$ , \*\* $p < .05$ , \*\*\* $p < .01$ .

**Table A7:** Faith-based facility ownership and consultation time and activities, controlling for caseload.

|  | (1)<br>Consultation time | (2)<br>Physical exam | (3)<br>Diagnostics | (4)<br>Drug prescribing |
| --- | --- | --- | --- | --- |
| FBP | 0.964***<br>(0.363) | 0.110***<br>(0.023) | 0.064***<br>(0.017) | 0.224***<br>(0.032) |
| Observation day 2 | 0.319*<br>(0.182) | 0.025*<br>(0.013) | -0.015*<br>(0.008) | 0.033*<br>(0.018) |
| Observation day 3+ | -0.208<br>(0.172) | -0.015<br>(0.011) | -0.029***<br>(0.008) | 0.080***<br>(0.017) |
| Observed during whole shift | 0.652***<br>(0.252) | 0.006<br>(0.022) | 0.022**<br>(0.009) | 0.113***<br>(0.030) |
| Survey month: February | 1.007**<br>(0.482) | -0.033<br>(0.035) | 0.041*<br>(0.023) | 0.059<br>(0.045) |
| Survey month: March | 0.566<br>(0.745) | 0.025<br>(0.046) | 0.055<br>(0.035) | -0.110*<br>(0.060) |
| Survey month: April | -0.738<br>(0.821) | -0.110**<br>(0.050) | 0.057<br>(0.041) | 0.087<br>(0.069) |
| Normal working day | -0.129<br>(0.143) | 0.013<br>(0.010) | -0.032***<br>(0.007) | 0.030**<br>(0.015) |
| Facility location: Rural | -0.209<br>(0.321) | 0.018<br>(0.021) | 0.045***<br>(0.016) | 0.169***<br>(0.030) |
| Facility type: Community Hospital | 2.627***<br>(0.502) | 0.087***<br>(0.029) | -0.027<br>(0.024) | -0.155***<br>(0.036) |
| Facility catchment population (1000) | 0.024***<br>(0.004) | 0.001***<br>(0.000) | 0.001***<br>(0.000) | 0.002***<br>(0.000) |
| Male healthcare worker | -0.020<br>(0.317) | 0.038**<br>(0.018) | -0.040**<br>(0.016) | -0.065**<br>(0.027) |
| Medical Assistant | -0.010<br>(0.270) | 0.011<br>(0.018) | 0.025**<br>(0.011) | -0.189***<br>(0.025) |
| Nurse Officer/Midwife Technician | 1.226*<br>(0.722) | 0.011<br>(0.041) | 0.061*<br>(0.037) | 0.341***<br>(0.046) |
| Professional experience (years) | 0.062***<br>(0.020) | 0.006***<br>(0.002) | -0.002*<br>(0.001) | -0.001<br>(0.002) |
| Health condition: ALRI | 0.384***<br>(0.145) | 0.059***<br>(0.012) | -0.021**<br>(0.008) | 0.031<br>(0.019) |
| Health condition: ANC | -3.107***<br>(0.743) | 0.442***<br>(0.062) | -0.049<br>(0.051) | -0.342***<br>(0.069) |
| Health condition: Diarrhoea | 0.877**<br>(0.386) | 0.062***<br>(0.023) | 0.034<br>(0.022) | -0.046<br>(0.036) |
| Health condition: Maternal/Newborn healthcare | 1.287<br>(0.806) | 0.046<br>(0.052) | -0.031<br>(0.046) | -0.211***<br>(0.056) |
| Health condition: NCDs | 2.170***<br>(0.313) | 0.075***<br>(0.019) | -0.021*<br>(0.011) | -0.071***<br>(0.024) |
| Health condition: Other | 1.442***<br>(0.188) | 0.089***<br>(0.014) | -0.032***<br>(0.008) | -0.068***<br>(0.020) |
| Patient order | -0.003**<br>(0.001) | -0.000***<br>(0.000) | -0.000<br>(0.000) | 0.000***<br>(0.000) |
| Number of patients seen/shift | -0.020***<br>(0.002) | -0.000***<br>(0.000) | -0.000**<br>(0.000) | 0.001***<br>(0.000) |
| Constant | -0.170<br>(0.778) | -0.066<br>(0.055) | -0.030<br>(0.032) | 0.077<br>(0.073) |
| Observations | 3905 | 3905 | 3905 | 3905 |
| R<sup>2</sup> | 0.167 | 0.161 | 0.088 | 0.231 |

*Notes:* This table presents the results from Equation 1, using data on consultations from healthcare-worker observations in outpatient departments in community hospitals and health centres participating in the Thanzi La Mawa (TLM) Time and Motion Study. The outcome variables are consultation time (minutes) and contents of the observed consultation. FBP equals 1 if the observed consultation was conducted at a faith-based (CHAM) facility and 0 if conducted at a government facility. The omitted reference category for observation day is Observation Day 1, for facility type it is health centre, for healthcare-worker profession it is Clinical Officer/Technician; and for health condition it is malaria. ALRI = acute lower respiratory infection, ANC= antenatal care, NCD = non-communicable disease. Robust standard errors are reported in parentheses. \* $p < .10$ , \*\* $p < .05$ , \*\*\* $p < .01$ .

**Table A8:** Robustness: Care received during outpatient consultation reported by patients.

|  | (1)<br>Physical examination | (2)<br>Any test | (3)<br>Diagnosed with new condition | (4)<br>Any treatment or medication |
| --- | --- | --- | --- | --- |
| FBP | 0.083<br>(0.078) | 0.135***<br>(0.034) | 0.140***<br>(0.027) | -0.031<br>(0.019) |
| Facility location: Rural | -0.229***<br>(0.071) | -0.051<br>(0.049) | 0.034<br>(0.048) | 0.048<br>(0.037) |
| Facility type: Community Hospital | 0.074<br>(0.058) | -0.052<br>(0.050) | 0.018<br>(0.046) | 0.013<br>(0.032) |
| Facility catchment population (1000) | -0.002***<br>(0.001) | 0.000<br>(0.000) | 0.002***<br>(0.000) | -0.001**<br>(0.000) |
| Patient age: 6–12 | -0.101**<br>(0.038) | -0.012<br>(0.054) | 0.030<br>(0.036) | 0.016<br>(0.044) |
| Patient age: 13–17 | -0.089<br>(0.068) | -0.018<br>(0.047) | 0.021<br>(0.044) | 0.074*<br>(0.035) |
| Patient age: 18–25 | -0.039<br>(0.066) | 0.053<br>(0.076) | 0.039<br>(0.033) | 0.027<br>(0.049) |
| Patient age: 26–49 | -0.098<br>(0.076) | -0.018<br>(0.073) | 0.067<br>(0.046) | 0.044<br>(0.048) |
| Patient age: 50+ | -0.088<br>(0.110) | 0.023<br>(0.066) | 0.147*<br>(0.077) | 0.075<br>(0.045) |
| Male Patient | -0.042<br>(0.029) | -0.042<br>(0.028) | 0.001<br>(0.021) | 0.018<br>(0.016) |
| Patient sought care for self | -0.025<br>(0.063) | -0.086<br>(0.055) | 0.033<br>(0.030) | -0.004<br>(0.033) |
| Household wealth score | 0.012<br>(0.012) | 0.009<br>(0.007) | 0.007<br>(0.005) | 0.003<br>(0.005) |
| Health condition: ALRI | -0.070<br>(0.089) | -0.469***<br>(0.082) | -0.009<br>(0.036) | -0.117**<br>(0.046) |
| Health condition: Diarrhoea | -0.256***<br>(0.059) | -0.464***<br>(0.137) | -0.022<br>(0.072) | -0.083<br>(0.051) |
| Health condition: NCDs | 0.078<br>(0.074) | -0.506***<br>(0.069) | 0.062<br>(0.072) | -0.130**<br>(0.058) |
| Health condition: Other | -0.017<br>(0.067) | -0.445***<br>(0.034) | -0.029<br>(0.031) | -0.107***<br>(0.019) |
| Health status: Mild symptoms | 0.429***<br>(0.133) | 0.466***<br>(0.133) | 0.051<br>(0.053) | 0.032<br>(0.083) |
| Health status: Moderate symptoms | 0.468***<br>(0.136) | 0.419***<br>(0.142) | 0.077<br>(0.047) | 0.034<br>(0.093) |
| Health status: Severe/Life-threatening symptoms | 0.475***<br>(0.151) | 0.428**<br>(0.156) | 0.185*<br>(0.095) | -0.017<br>(0.110) |
| Constant | 0.280<br>(0.187) | 0.570***<br>(0.150) | -0.166***<br>(0.040) | 0.936***<br>(0.096) |
| Observations | 1118 | 1118 | 1118 | 1118 |
| $R^2$ | | | | |

*Notes:* This table presents the results from Equation 1, using data from the patient-exit interviews conducted as part of the Thanzi La Mawa (TLM) project with patients leaving community hospitals and health centres who reported attending an outpatient department. The outcome variables capture the likelihood that a patient reported receiving a physical examination, any diagnostic test, a new diagnosis, or any treatment/medication during their visit. FBP equals 1 if the patient attended a consultation at a faith-based (CHAM) facility and 0 if they visited a government facility. The omitted reference category for facility type is health centre, for patient age is 0–5 years, and for health condition is malaria. ALRI = acute lower respiratory infection; NCD = non-communicable disease. Robust standard errors in parentheses. Standard errors and significance stars for the coefficient on FBP are based on wild cluster bootstrap (999 replications). \* $p < .10$ , \*\* $p < .05$ , \*\*\* $p < .01$ .

## Footnotes

1 The data collection is described further in Nkhoma, et al. (2024).

2 Bachelor of Medicine and Bachelor of Surgery (MBBS), a Bachelor of Nursing, Bachelor of Health Management or related qualifications.

3 The data collection tool originally included 127 pre-recorded activity codes developed in collaboration with Malawian clinical experts. If enumerators were not able to identify an appropriate existing activity code, they provided further text information about the activity. The text data were later reviewed and re-categorised into the previously established activity codes or organised into new frequently observed activity categories resulting in 167 activity categories.

4 1.51% of all patients in our sample returned to the same healthcare worker per shift. Given that we do not know whether a patient may have returned to a different healthcare worker during the same day, we follow previous studies (see Irving et al. (2017)) and focus on the duration of the initial consultation only and remove return consultations within the same day.

5 Very few clients reported their health condition or reason for seeking care to be related to ANC and maternal and newborn care. Therefore, these conditions are included in “Health condition: Other”.

6 We do not include survey-month covariates because of the limited sample size and their collinearity with facility ownership and facility type.

## Notes

### Competing Interest Statement

The authors have declared no competing interest.

### Funding Statement

This study was funded by The Wellcome Trust (Thanzi La Mawa; reference 223120/Z/21/Z).

### Author Declarations

College of Medicine Research Ethics Committee of Kamuzu University of Health Sciences gave ethical approval for this work.

